# Prevalence of Relapse in Pulmonary Sarcoidosis: A Systematic Review and Meta-analysis

**DOI:** 10.1101/2025.07.15.25330996

**Authors:** Zia Hashim, Robert P Baughman, Ritesh Agarwal, Naresh Kumar Tripathy, Mansi Gupta, Ajmal Khan, Ajit Kumar, Anup Kumar, Alok Nath

**Affiliations:** Department of Pulmonary Medicine, Sanjay Gandhi Postgraduate Institute of Medical Sciences, Lucknow, India; Department of Internal Medicine, University of Cincinnati Medical Center, Cincinnati, OH, USA; Department of Pulmonary Medicine, Postgraduate Institute of Medical Education and Research, Chandigarh, India; Department of Hematology, Sanjay Gandhi Postgraduate Institute of Medical Sciences, Lucknow, India; Department of Biostatistics and Health Informatics, Sanjay Gandhi Postgraduate Institute of Medical Sciences, Lucknow, India

**Author notes:** Zia Hashim and Alok Nath are joint corresponding authors Correspondence: Zia Hashim, SGPGIMS, Lucknow, India.

**Keywords:** pulmonary sarcoidosis, relapse, prevalence, risk factors, meta-analysis, heterogeneity

## Abstract

**Background and objective:** Relapse after treatment discontinuation or reduction frequently occurs in pulmonary sarcoidosis (PS), but reported rates vary widely. We conducted a systematic review and meta-analysis to estimate relapse prevalence and evaluate risk factors.

**Methods:** We searched PubMed, Embase, Scopus, and Google Scholar from inception through January 2026. Two reviewers independently extracted data on relapse prevalence, timing, definitions, and predictors, and assessed study quality using the Hoy et al. tool. We calculated pooled prevalence using random effects models, assessed heterogeneity using I², and performed subgroup analyses and meta-regression.

**Results:** We included 51 studies comprising 6093 patients; 3682 were followed up, and 1442 relapsed. Pooled relapse prevalence was 0.39 (95% CI: 0.33–0.45) with substantial heterogeneity (I²=94%). Relapse prevalence was lower in prospective (0.31, 95% CI: 0.25–0.37) than retrospective (0.45, 95% CI: 0.36–0.54) studies. Only 23 (45%) studies provided explicit relapse definitions. Advanced radiographic stage was associated with higher relapse odds than stage I (log OR 0.55–1.04, corresponding OR 1.73–2.83; p<0.05), and Black race with higher odds than White race (log OR 0.91, corresponding OR 2.48; p<0.01).

**Conclusions:** Approximately 39% of PS patients relapse after treatment reduction or discontinuation. Marked heterogeneity in relapse definitions underscores the need for standardized criteria. Clinical assessment appears comparable to complex definitions for assessing relapse.

**Funding:** None.

**Registration:** PROSPERO CRD420251075384.

**HIGHLIGHS:**

- First meta-analysis of relapse prevalence in pulmonary sarcoidosis
- Pooled relapse prevalence was 39% (51 studies, 6093 patients)
- Prospective studies showed lower relapse rates (31%) than retrospective (45%)
- Only 45% of studies provided explicit definitions of relapse
- Advanced radiographic stage and Black race predicted higher relapse risk

## INTRODUCTION

Sarcoidosis is a multisystem disorder characterized histologically by non-caseating granulomas, most commonly affecting the lungs and intrathoracic lymph nodes [1, 2]. The clinical course of pulmonary sarcoidosis (PS) is highly variable. While approximately one-third of patients experience spontaneous remission within 2–3 years of diagnosis, a significant proportion develop persistent or relapsing disease requiring prolonged immunosuppression. Historically, systemic glucocorticoids remain the cornerstone of therapy for clinically significant PS. Current practice commonly involves initiating prednisone around 20 mg daily, followed by a gradual taper over 6 months guided by clinical and radiological response [3]. However, methotrexate can also be used as a first-line therapy, as demonstrated by the recently published PREDMETH trial [4]. Yet, a vexing problem in sarcoidosis management is determining the optimal duration of therapy as prolonged therapy increases the risk of cumulative toxicity, while premature reduction or discontinuation can lead to disease relapse.

Relapse in PS, broadly defined as worsening of clinical, radiological, or functional status after treatment discontinuation or reduction, represents a major therapeutic challenge. Relapse is associated with impaired quality of life, increased healthcare utilization, and potential risk of progressive pulmonary fibrosis. Despite its importance, published estimates of relapse rates vary widely, ranging from 13% to 80% in previous reviews [5, 6]. Inconsistent definitions of relapse, including varying combinations of clinical, functional, radiological, and biomarker parameters, may yield different relapse rates across studies.

To address these gaps, we conducted a systematic review and meta-analysis with the primary objective of estimating the pooled relapse prevalence in patients with PS following treatment reduction or discontinuation. Secondary objectives were: (1) to describe the timing and patterns of relapse events; (2) to identify demographic, clinical, radiographic, and treatment factors associated with relapse risk; (3) to evaluate the variability in relapse definitions across studies; and (4) to assess the influence of study design and methodological quality on relapse prevalence estimates.

## METHODS

### Study protocol and registration

We conducted this systematic review and meta-analysis in accordance with the PRISMA and MOOSE guidelines [7, 8]. We registered the study protocol prospectively with PROSPERO (CRD420251075384). Ethics committee approval was not required, as this was an analysis of published data.

### Search strategy

We systematically searched PubMed, Embase, Scopus, Google Scholar, ClinicalTrials.gov, and PROSPERO from database inception through January 31, 2026. We used a search strategy combining Medical Subject Headings and Emtree terms with relevant free-text keywords linked by Boolean operators (Supplementary material). We supplemented the electronic search with manual screening of reference lists, forward citation tracking, and gray literature searches including abstracts from ATS and ERS annual conferences (2010–2025).

### Study selection

We imported search results into EndNote 25 (Clarivate Analytics, Philadelphia, PA) to remove duplicates and screen them. Two authors (Z.H. and N.K.T.) independently screened titles and abstracts, followed by full-text evaluation of potentially eligible articles. Discrepancies were resolved through consensus discussion, with the help of a third reviewer (A.K.) when necessary. When relapse data were incomplete or full texts were unavailable, we contacted the corresponding authors.

### Eligibility criteria

Inclusion: Studies were included if they: (1) included patients with biopsy-proven or clinically diagnosed sarcoidosis; and (2) reported relapse rates following treatment discontinuation or dose reduction, with explicit or derivable definitions.

Exclusion: Studies were excluded if they: (1) enrolled fewer than 10 patients; (2) reported isolated extrapulmonary sarcoidosis without pulmonary involvement; (3) had inadequate or unclear definitions of disease states; (4) were inaccessible despite author contact; or (5) had overlapping cohorts (in which case the most recent or comprehensive publication was included).

### Definitions of sarcoidosis disease states

Early relapse: worsening of disease activity during or within 1 month of treatment tapering or withdrawal, without a documented interval of sustained remission. The 1-month boundary adopted here is operational and pragmatic rather than evidence-derived.

Late relapse: recurrence of disease activity following a documented period of sustained remission (≥1 month), manifested by reappearance of clinical symptoms, new or worsening radiological abnormalities, functional decline, or elevation of biomarkers — representing true disease recurrence after a phase of immunological inactivity.

Remission: a disease-free state persisting for ≥1 month, characterized by absence of clinical symptoms, stable or improved radiological findings, stable or improved pulmonary function, and normalized or stable biomarkers.

### Data extraction

Two reviewers (Z.H. and N.K.T.) independently extracted data using a piloted standardized form: (1) study characteristics (first author, year, country, design); (2) patient demographics (sample size, age, sex, ethnicity); (3) clinical characteristics (organ involvement, Scadding stage); (4) treatment details (type and duration); and (5) outcome data (frequency of relapse, relapse definitions, time to relapse). For risk factor analysis, we extracted stage-specific relapse rates and demographic-stratified outcomes.

### Assessment of study quality

Methodological quality was independently assessed using the Hoy et al. (2012) risk of bias tool for prevalence studies [9]. Studies with total scores of 0–2 were classified as low risk of bias, 3–4 as moderate risk, and 5–10 as high risk of bias.

### Statistical analysis

#### Pooled prevalence estimation

We pooled relapse proportions using a random-effects model (restricted maximal likelihood method), reporting untransformed proportions with 95% confidence intervals [10].

#### Publication bias

We assessed publication bias through funnel plot inspection and Egger’s regression test and Begg’s rank correlation test (p<0.05 indicating potential bias) [11, 12].

#### Sensitivity analyses

We performed multiple sensitivity analyses: (1) exclusion of outliers by Baujat and Galbraith plots; (2) leave-one-out analysis; (3) Freeman-Tukey transformation; (4) restriction to studies with explicit relapse definitions; (5) exclusion of small studies (≤50 patients); (6) exclusion of paediatric populations; (7) restriction to low-risk-of-bias studies; and (8) Bayesian hierarchical random-effects model.

#### Heterogeneity assessment

Statistical heterogeneity was quantified using Higgins’ I² statistic (I²≥50% indicating substantial heterogeneity) and Cochran’s Q test (p<0.10) [13]. Baujat and Galbraith plots were constructed to identify outliers [14, 15].

#### Meta-regression and subgroup analyses

We performed multivariate meta-regression using a restricted maximum-likelihood random-effects model. Covariates included: (1) study design; (2) explicit relapse definition; (3) publication era (before/after 2000); (4) geographic region (Asian vs. non-Asian); (5) study quality; and (6) sample size. Univariate subgroup analyses used Cochran’s Q test.

#### Risk factor analysis

Binary risk factors (age, sex, radiographic stage) were expressed as log odds ratios (LORs) with 95% CIs. Continuous variables used pooled mean differences with 95% CIs from inverse-variance random-effects models. OR = e^LOR. Predictors examined: age; sex; race (Black vs. White); radiographic stage (stage IV vs. I; stages II–III vs. I; stages II–III vs. IV).

#### Duration of therapy

Treatment duration was standardized to months. For studies reporting duration by outcome group, a weighted mean was calculated; for those providing only ranges, midpoint approximations were used. Studies with no estimable duration were coded ‘NR’ and excluded from duration-based meta-regression (21/51 studies, 41%). An 18-month cutoff was predetermined, aligning with ERS/ATS consensus recommendations [5].

#### Certainty of evidence

We assessed the certainty of evidence using the GRADE framework, adapted for prevalence studies, considering risk of bias, inconsistency, indirectness, imprecision, and publication bias [16].

## RESULTS

### Study selection

Our systematic search identified 1972 records (1960 from databases and 12 from registers). After removing 650 duplicates, we screened 1,322 articles and identified 85 studies for full-text assessment. Of these, 34 were excluded. We included 51 studies comprising 6093 patients (Figure 1, Table 1).

**Figure 1.**
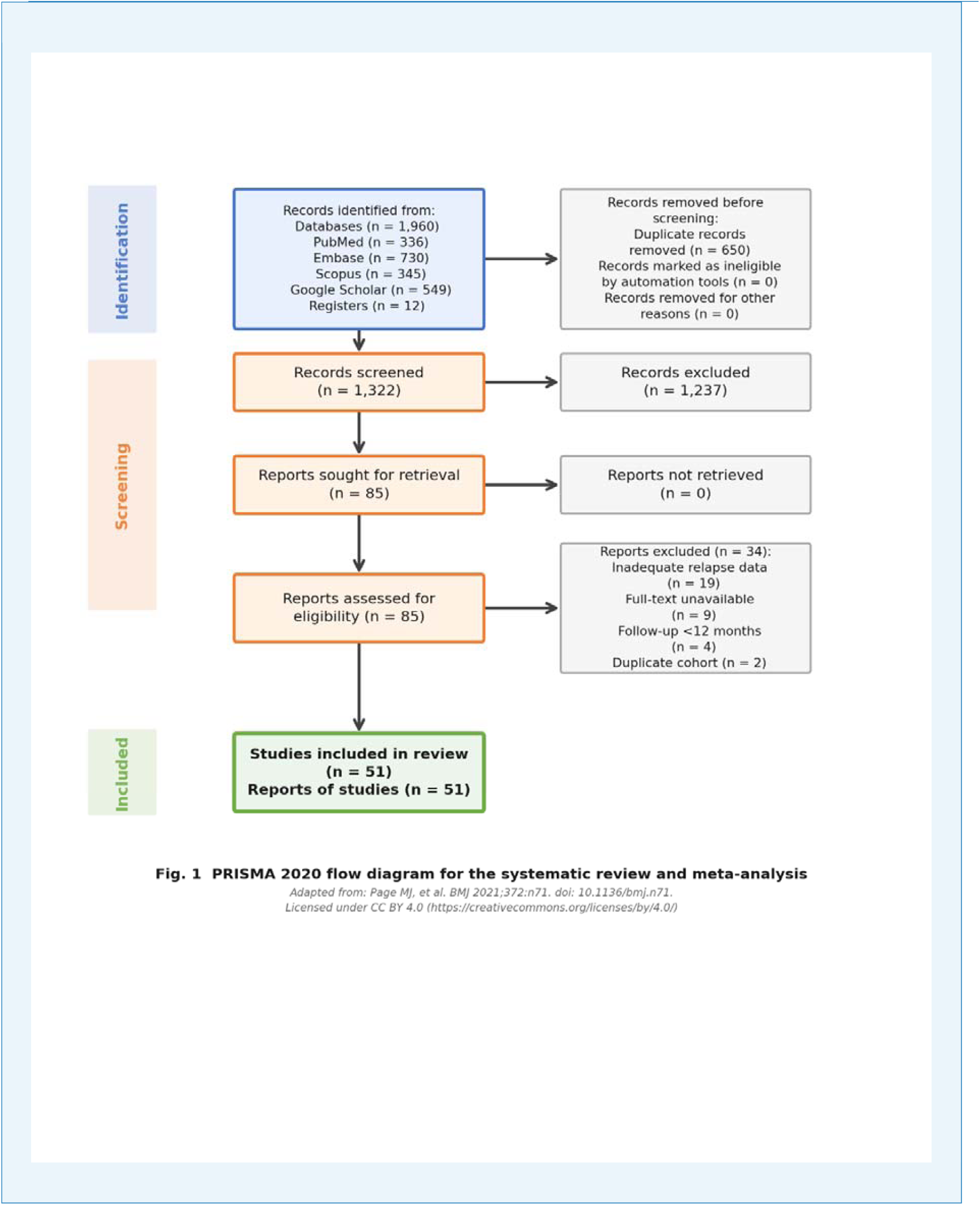
PRISMA flow diagram depicting the study selection process, including identification, screening, eligibility assessment, and inclusion of eligible studies.

**Table 1.**
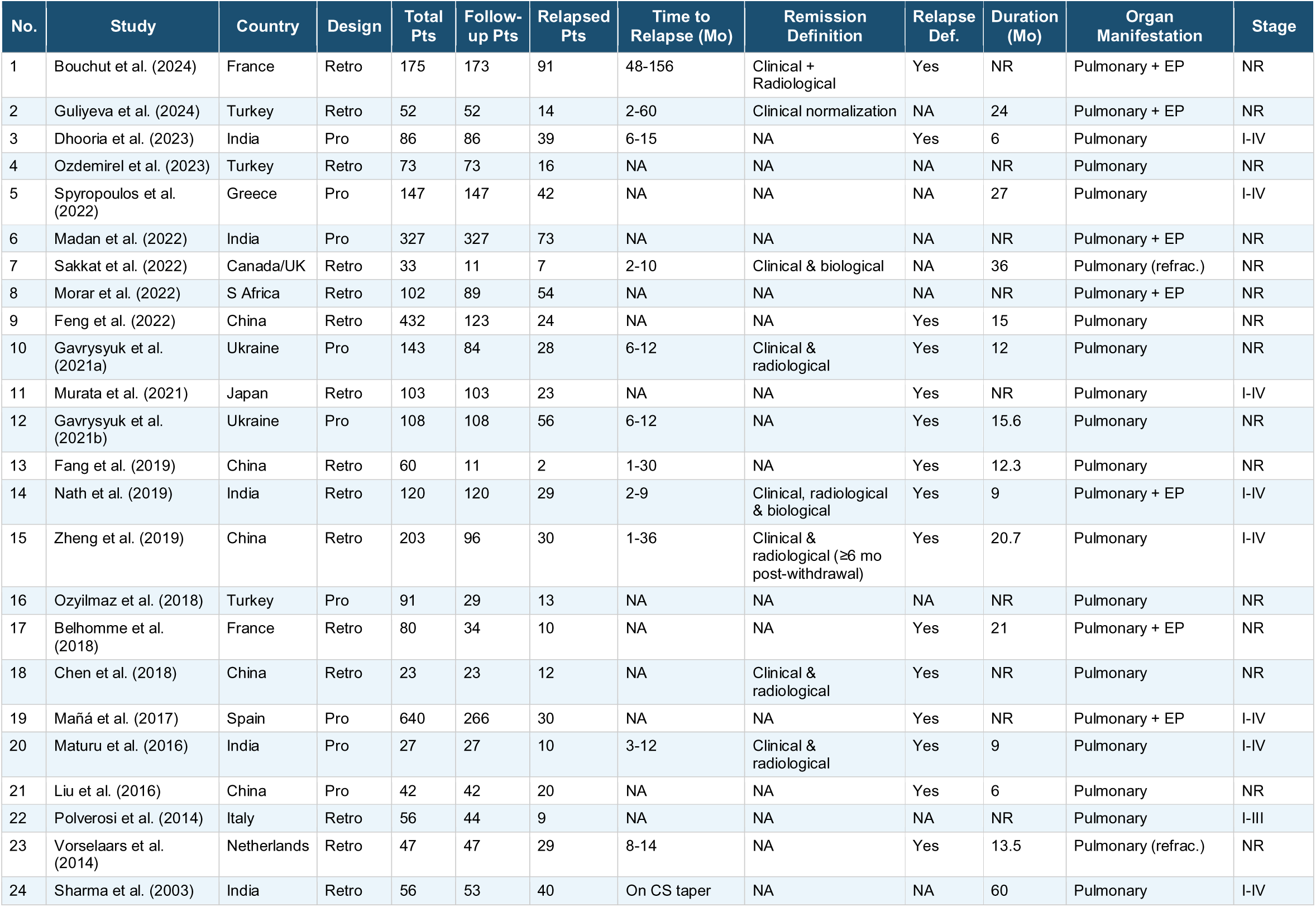
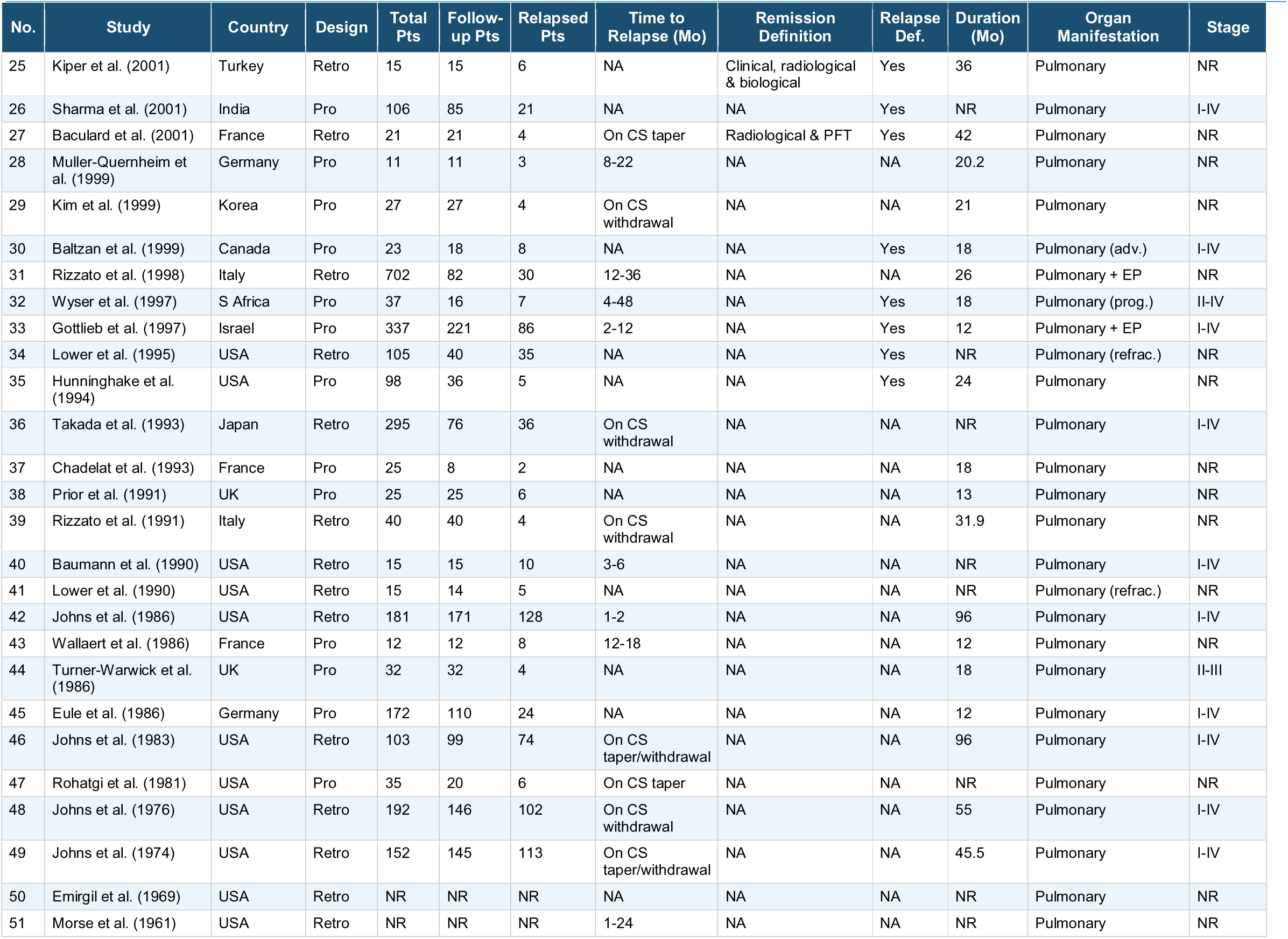
Studies included in the systematic review and meta-analysis. Pro = prospective; Retro = retrospective; EP = extrapulmonary; CS = corticosteroid; NR = not reported; NA = not available; Mo = months; adv. = advanced; prog. = progressive; refrac. = refractory.

### Study characteristics

Studies were published between 1961 and January 2026, with 27 (53%) published after 2000. Most were from non-Asian countries (33 studies, 65%). Twenty-two (43%) were prospective and 29 (57%) were retrospective. The median sample size was 44 patients (IQR 21.5–101); follow-up ranged from 6 months to 40 years (Table 1).

### Methodological quality

Thirty-four (67%) studies were classified as low risk of bias and 17 (33%) as moderate risk; none was judged as high risk of bias (Figure S1). Sources of potential bias included methodological limitations (52%), inadequate sample size (50%), concerns about measurement validity (48%), unclear study descriptions (26%), and measurement reliability issues (12%). There was no difference in relapse prevalence between low- and moderate-risk-of-bias studies (Figure S2).

### Primary objective: overall prevalence of relapse

Among 6093 patients, 3682 were followed up and 1442 relapse events were reported. The pooled relapse prevalence was 0.39 (95% CI: 0.33–0.45), with individual study estimates ranging from 0.10 to 0.88. We observed substantial statistical heterogeneity (I²=94%, τ²=0.04, Cochran’s Q=1044, p<0.01) (Figure 2). Prospective studies showed a significantly lower pooled relapse prevalence of 0.31 (95% CI: 0.25–0.37) compared with retrospective studies (0.45, 95% CI: 0.36–0.54) (p=0.01) (Table 2, Figure S3).

**Figure 2.**
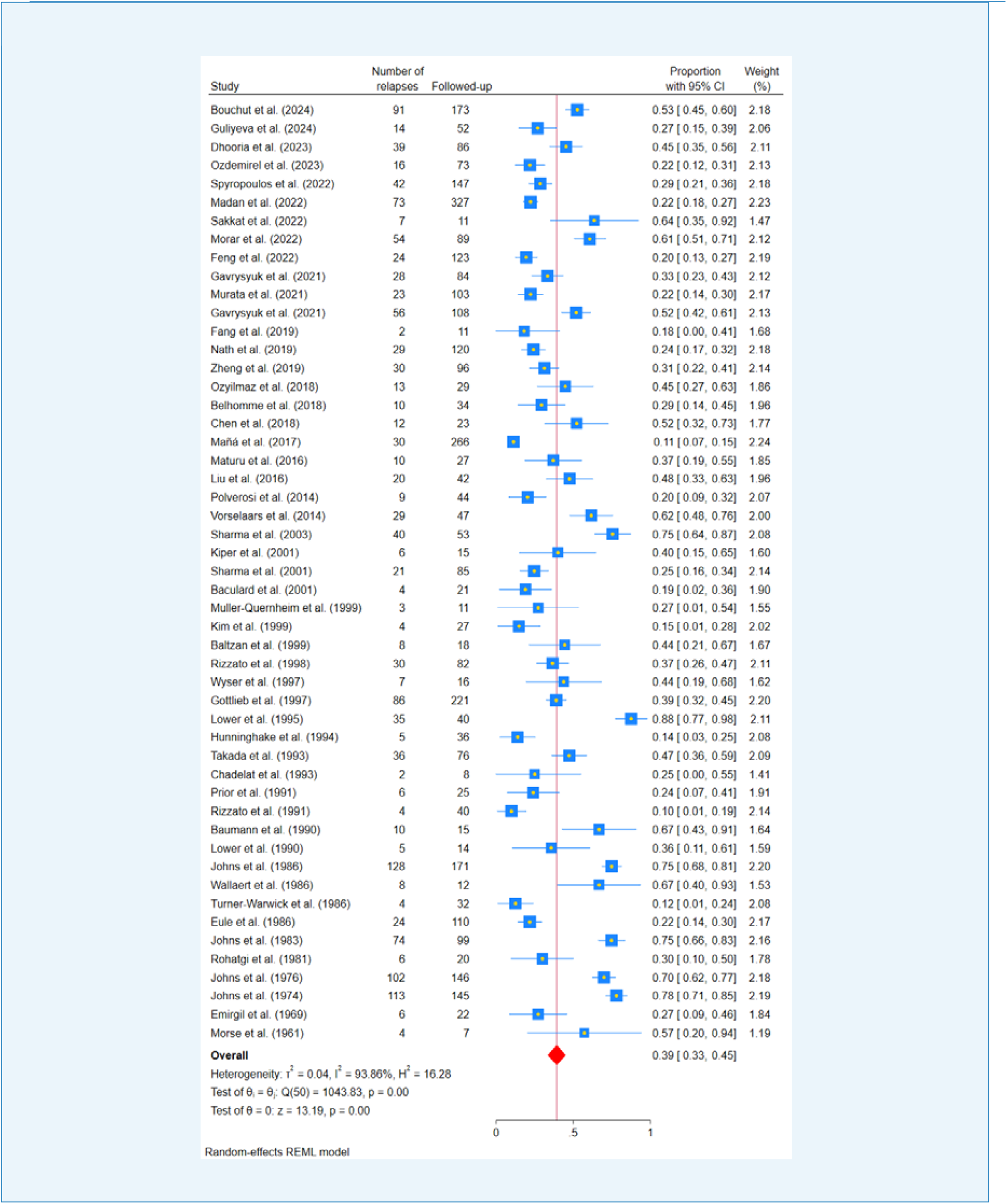
Forest plot of pooled relapse prevalence in pulmonary sarcoidosis. Individual study estimates (blue squares) and 95% confidence intervals (horizontal lines) are shown, with the overall pooled estimate (red diamond) of 0.39 (95% CI: 0.33–0.45) derived from a random-effects REML model. Substantial heterogeneity was present (I²=94%).

**Table 2.** Summary of sensitivity analyses and subgroup analyses of pooled relapse prevalence in pulmonary sarcoidosis.

| Analysis Type / Subgroup | N (studies) | Pooled Prevalence (95% CI) | I<sup>2</sup> (%) | τ<sup>2</sup> |
| --- | --- | --- | --- | --- |
| **Primary Analysis** |  |  |  |  |
| Overall (all studies) | 51 | 0.39 (0.33–0.45) | 94 | 0.04 |
| **Study Design** |  |  |  |  |
| Prospective studies | 22 | 0.31 (0.25–0.37) | 86 | 0.01 |
| Retrospective studies | 29 | 0.45 (0.36–0.54) | 97 | 0.04 |
| **Outlier Analysis** |  |  |  |  |
| Removing 10 outlier studies | 41 | 0.35 (0.31–0.40) | 82 | 0.01 |
| **Study Quality** |  |  |  |  |
| Low-risk-of-bias (high-quality) studies | 34 | 0.41 (0.34–0.48) | 97 | 0.04 |
| **Definition Consistency** |  |  |  |  |
| Studies with explicit relapse definition | 23 | 0.37 (0.29–0.44) | 92 | 0.03 |
| **Population Homogeneity** |  |  |  |  |
| Adults only | 47 | 0.40 (0.34–0.46) | 94 | 0.04 |
| **Sample Size** |  |  |  |  |
| Studies with >50 follow-up patients | 24 | 0.41 (0.33–0.50) | 97 | 0.04 |
| **Statistical Methods** |  |  |  |  |
| Freeman-Tukey double arcsine transformation | 51 | 0.39 (0.32–0.45)* | 92 | 0.17 |
| Bayesian hierarchical random-effects model | 51 | 0.39 (0.32–0.45) | 99 | 0.08 |
| **Treatment Duration†** |  |  |  |  |
| Corticosteroids ≤18 months | 17 | 0.35 (0.28–0.43) | 84 | 0.02 |
| Corticosteroids >18 months | 13 | 0.26 (0.20–0.32) | 62 | 0.01 |
| **Leave-one-out Analysis** |  |  |  |  |
| Range across all leave-one-out iterations | 51 | 0.40 (0.33–0.46) | 94 | 0.04 |
| * Reported as back-transformed proportion. † Restricted to 30 studies with estimable treatment duration data. ES = effect size (pooled prevalence); CI = confidence interval; I<sup>2</sup> = heterogeneity index; τ<sup>2</sup> = between-study variance. |  |  |  |  |

Multiple sensitivity analyses demonstrated the robustness of the pooled estimate. Exclusion of 10 outlier studies based on Baujat and Galbraith plots reduced heterogeneity by 12% (I²=82%) with minimal change in the pooled estimate (0.35, 95% CI: 0.31–0.40) (Table 2, Table S1–S2, Figures S3–S9).

### Secondary objectives

#### Timing of relapse

Nineteen (37%) studies reported time-to-relapse data (Table 1, Table S5, Figure S10). The median time-to-first late relapse was 10.5 months (IQR 6.5–17.0 months; range 1–156 months). A sensitivity analysis adjusting for left-truncation bias yielded a median of 8.2 months (IQR 4.5–15.0 months). Sixty-three percent of studies (12/19) reported relapses within 1.5 years, and 84% (16/19) within 3 years. Pooled proportions ranged from 0.431 to 0.469 across all boundary cutoffs (≥1, ≥2, ≥3, ≥6 months), with fully overlapping 95% CIs.

#### Demographic characteristics

Age was not associated with relapse (mean difference: −0.05 years, 95% CI: −1.56 to 1.47; p=0.95; 15 studies, n=1,434). Female sex was not significantly associated (LOR: 0.19, 95% CI: −0.11 to 0.49; p=0.21; 15 studies, n=1,326). Black patients had significantly higher odds of relapse than White patients (LOR: 0.91, 95% CI: 0.44–1.37; OR: 2.48, 95% CI: 1.55–3.94; p<0.01; 5 studies, n=370). Pooled prevalence in adult-only studies (47 studies) was 0.40 (95% CI: 0.34–0.46) (Figure S14).

#### Radiographic staging

Compared with Scadding stage I disease (reference), the odds of relapse were significantly higher in stage IV (LOR: 0.55, 95% CI: 0.07–1.03; OR: 1.73, 95% CI: 1.07–2.80; p=0.02) and in stages II–III combined (LOR: 1.04, 95% CI: 0.25–1.83; OR: 2.83, 95% CI: 1.28–6.23; p=0.01). No significant difference was observed between stages II–III and stage IV (LOR: −0.54, 95% CI: −1.39 to 0.31; p=0.21) (Figure S15).

#### Disease extent

Relapse prevalence did not differ significantly between isolated pulmonary sarcoidosis and multiorgan involvement (0.39, 95% CI: 0.33–0.45; p=0.66) (Figure S16).

#### Treatment regimens

Corticosteroids alone were used in 37 (73%) studies, steroid-sparing agents in 7 (14%), and combination therapy in 7 (14%) studies. Combination therapy showed a numerically lower relapse prevalence (0.31, 95% CI: 0.21–0.41) compared with corticosteroids alone (0.38, 95% CI: 0.32–0.45) or steroid-sparing agents alone (0.54, 95% CI: 0.36–0.72), but differences were not statistically significant (p=0.08) (Figure S17).

#### Duration of therapy

Studies with corticosteroid duration ≤18 months (17 studies, n=1,090) had higher pooled relapse prevalence (0.35; 95% CI: 0.28–0.43; I²=84%) compared with those >18 months (13 studies, n=594) (0.26; 95% CI: 0.20–0.32; I²=62%). The between-subgroup difference approached but did not reach statistical significance (Cochran Q=3.82, p=0.05) (Figure S18).

#### Geographic and temporal factors

No significant differences in relapse prevalence were observed between Asian and non-Asian studies (0.34 vs. 0.42; p=0.12) or between studies published before and after 2000 (0.43 vs. 0.36; p=0.24) (Figure S3).

### Heterogeneity in relapse definitions

Only 23 studies (45%) provided explicit definitions of relapse (Table 1, Table S4). Definitions varied: clinical criteria alone (9 studies, 39%) and composite criteria (14 studies, 61%). There was no significant difference in relapse prevalence between clinical-criteria-only and composite-criteria studies (0.37, 95% CI: 0.29–0.44, p=0.71) (Figure S19).

### Meta-regression

Multi-variable meta-regression incorporating six prespecified covariates found only study design as a significant predictor of relapse prevalence (coefficient=0.148, SE=0.060; p=0.014). A pre-specified univariate meta-regression with treatment duration as the sole predictor was also performed, restricted to the 30 studies with estimable duration data (Table S6).

### Publication bias

Visual inspection of the funnel plot revealed no asymmetry (Figure 3). Neither Egger’s test (intercept=0.50, 95% CI: −2.11 to 3.10; p=0.70) nor Begg’s test (Kendall’s τ=0.07; p=0.47) indicated publication bias.

**Figure 3.**
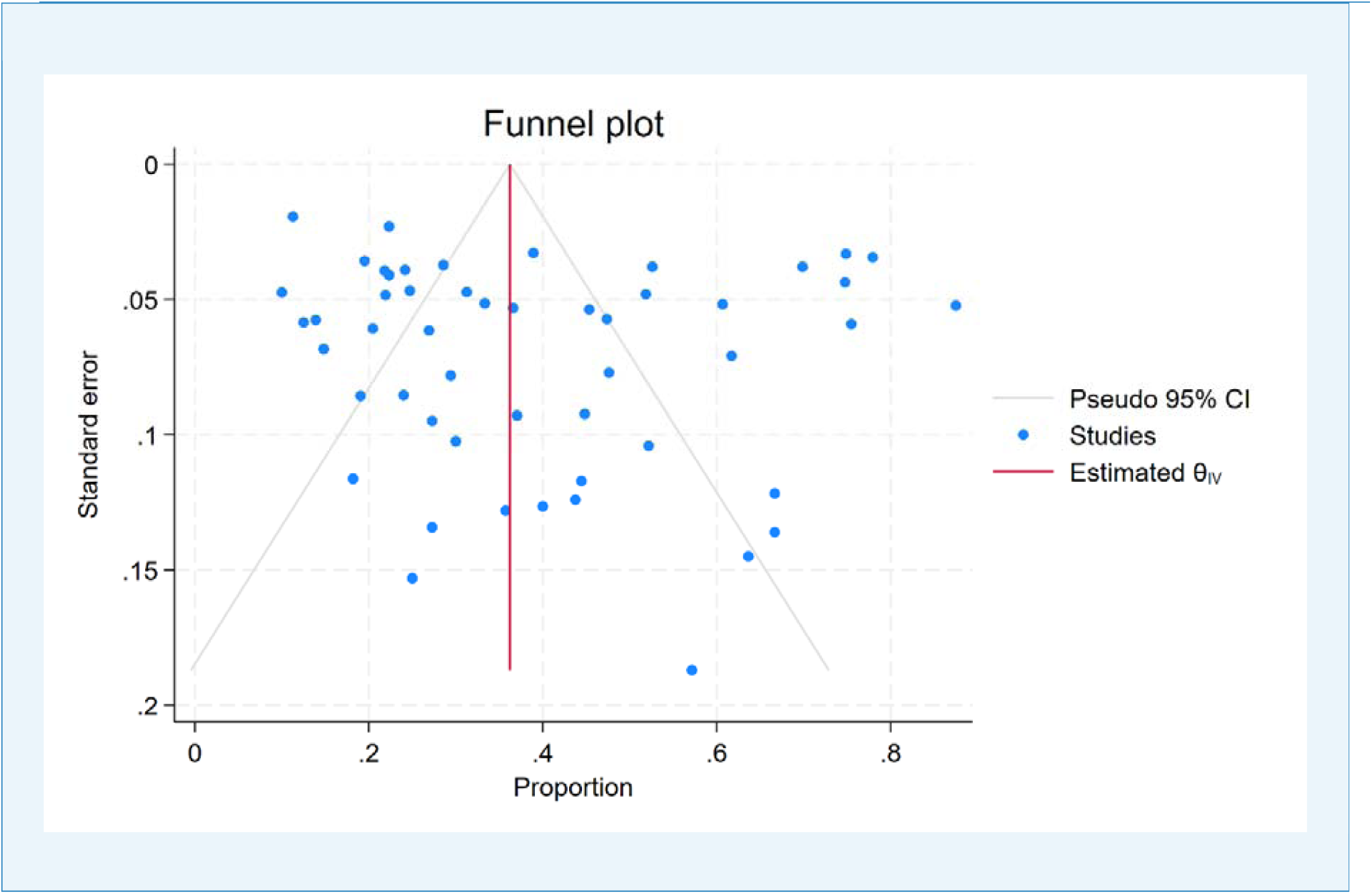
Funnel plot for publication bias. Symmetric distribution of studies around the pooled estimate suggests no publication bias (Egger’s p=0.70; Begg’s p=0.47).

### Certainty of evidence

Using GRADE methodology, we rated the overall certainty of evidence for relapse prevalence as low, downgraded for risk of bias and inconsistency. Evidence from prospective studies and the association between Black race and relapse was rated moderate certainty (Table S7).

## DISCUSSION

In this systematic review and meta-analysis of 51 studies comprising 6093 patients with PS, we found a pooled relapse prevalence of 39%. Individual study estimates varied widely (10–88%), reflecting marked variability in study designs, relapse definitions, and follow-up practices. Prospective cohorts reported a lower relapse prevalence (31%) than retrospective studies (45%). In addition, radiographic stages II–IV and Black race were significant predictors of relapse. To our knowledge, this study represents the first comprehensive assessment of relapse burden and associated predictors in patients with PS.

A key finding of our analysis was the substantial heterogeneity across studies (I²=94%). This heterogeneity reduced the certainty of evidence for the pooled relapse prevalence estimate (rated low by GRADE). Nevertheless, high heterogeneity is common in meta-analyses of observational data and does not invalidate results when thoroughly explored. Multiple sensitivity analyses demonstrated remarkable consistency, including leave-one-out analysis and Bayesian hierarchical modeling, which yielded prevalence estimates of 39–40%. Excluding outlier studies reduced heterogeneity modestly (I²=82%) with similar pooled prevalence, indicating that outlier studies contributed to, but do not fully explain, the observed variability. Despite prospective studies showing lower relapse rates overall, substantial heterogeneity persisted even within this subgroup (I²=86%), indicating that study design alone does not fully account for the observed variability.

Several factors likely contributed to heterogeneity. Patient populations varied significantly: certain prospective cohorts primarily consisted of Asian patients with stage II disease, a group experiencing frequent spontaneous remissions. On the other hand, some cohorts consisted of Black or White patients with more advanced, chronic illnesses, which are linked to an increased inherent risk of relapse. Organ involvement also varied; isolated pulmonary disease carries a different trajectory than multiorgan sarcoidosis, where cardiac, neurological, or cutaneous involvement typically signals a more refractory course. Treatment protocols were inconsistently reported, including the dose at which tapering began, the rate of reduction, and minimum treatment duration — all independently influencing relapse risk. Follow-up duration ranged from six months to several decades, and the studies spanned six decades of evolving diagnostic and therapeutic standards.

In multivariable meta-regression, study design was the only significant variable associated with relapse prevalence. Retrospective cohorts demonstrated higher pooled relapse prevalence than prospective cohorts, reflecting limitations including ascertainment bias, variable relapse definitions, incomplete follow-up, and inconsistent evaluation of competing diagnoses. Limiting the analysis to studies with explicit relapse definitions or high-quality studies resulted in only modest reductions in pooled relapse prevalence and did not meaningfully reduce heterogeneity. These findings suggest that variations in relapse prevalence are driven not solely by methodological quality but also by clinical differences across populations, treatment strategies, and follow-up durations.

A major limitation highlighted by our review was the lack of standardized definitions of relapse. Only 45% of the included studies provided explicit relapse criteria, with definitions varying widely across clinical, radiological, functional, biomarker-based, and composite domains. Subgroup analysis revealed no significant difference in relapse rates between studies using clinical criteria and those incorporating composite criteria. This supports the practical observation that relapse identification remains largely clinical, consistent with prior task force recommendations [17].

A fundamental conceptual challenge is the difficulty of distinguishing true disease recurrence from corticosteroid-dependent disease activity. When symptoms re-emerge during treatment or within a short window of its withdrawal, before any sustained period of immunological quiescence has been established, this pattern more accurately reflects treatment failure or non-remission than genuine relapse. Baughman and Judson [18] noted that many apparent relapses represent chronic disease that was suppressed rather than remitted, while Gottlieb et al. [19] required a sustained treatment-free interval exceeding one month before any recurrence could qualify as relapse. We adopted a one-month operational threshold to separate these two phenomena. Sensitivity analyses using cutoffs of ≥1, ≥2, ≥3, and ≥6 months produced pooled estimates of 0.43–0.47 with fully overlapping confidence intervals (Supplementary Table S5), confirming robustness regardless of where this boundary is drawn.

The timing of relapse has important implications for clinical care. Across studies, the majority (63%) occurred within 1.5 years of treatment discontinuation, with peak risk during months 6–12. Relapses were also observed late in a subset of studies, supporting the need for extended follow-up in patients with persistent disease risk or advanced pulmonary involvement.

Radiographic staging was an important predictor of relapse risk; stages II–IV were associated with significantly higher relapse odds compared with stage I disease. Interpretation of relapse in stage IV patients warrants caution because fibrotic lungs are vulnerable to infection and structural complications, and distinguishing true sarcoidosis relapse from infectious exacerbations or progressive fibrotic decline may be challenging. Future studies should incorporate standardized protocols for excluding alternative infectious and non-infectious causes [20].

We found no significant difference in relapse prevalence between isolated PS and multiorgan disease. However, prior studies suggest higher relapse rates in patients with multiorgan disease [21, 22], and in those with lung [23], heart [24], musculoskeletal system [25], eye [26], skin [26], and neurological involvement [27, 28]. Isolated erythema nodosum or peripheral lymphadenopathy was associated with a lower relapse risk [19, 29]. Prospective studies are needed to determine whether organ-specific involvement can be incorporated into relapse prediction models.

Black patients had significantly higher odds of relapse compared with White patients, consistent with well-established racial disparities in sarcoidosis severity. This heightened relapse risk likely reflects a combination of genetic predispositions [30], socioeconomic factors such as healthcare access and treatment adherence [31], or differences in immune responses [32]. There was no effect of age, sex, publication era, or geographic region on relapse prevalence.

Corticosteroids remain the traditional first-line treatment. In our subgroup analysis, relapse prevalence was not significantly different across treatment types, with comparable rates in corticosteroid monotherapy, steroid-sparing immunosuppressants alone, or combination therapy. However, the numerically lower relapse prevalence with combination therapy should be interpreted cautiously, as treatment selection is confounded by baseline disease severity. Recently, the PREDMETH trial found comparable outcomes between methotrexate and prednisone monotherapy in selected treatment-naïve symptomatic PS patients [4], and observational data suggest that steroid-sparing agents may reduce cumulative corticosteroid exposure [33].

Treatment duration emerged as a potentially important modulator of relapse risk. Patients on corticosteroids for 18 months or less had a 35% relapse rate, while those treated longer had a 26% relapse rate (p=0.05). Given the considerable heterogeneity in both subgroups (I²=84% and 62%), duration by itself is an insufficient explanation. Optimal treatment duration in pulmonary sarcoidosis remains undefined, and prospective trials with standardised protocols are long overdue.

Our study has several limitations. First, heterogeneity in relapse definitions was substantial, and more than half of the studies lacked explicit criteria. Second, the predominance of retrospective studies increased ascertainment and reporting biases. Third, reporting of treatment duration, tapering protocols, and remission depth was inconsistent. Fourth, geographic representation was skewed towards Europe and North America. Fifth, the data were insufficient to evaluate relapse predictors, such as biomarkers (serum ACE), PET activity, or genetic susceptibility. Finally, individual-patient data were unavailable for meta-analysis.

In conclusion, relapses occur in approximately 39% of patients with PS following treatment reduction or discontinuation, predominantly within the first two years. Advanced radiographic stage and Black race are associated with increased relapse risk, whereas age, sex, disease extent, publication era, and treatment modality are not. While clinical assessment remains a practical and widely applicable approach to monitoring relapse, the absence of standardized relapse criteria substantially limits certainty [35, 36]. Our findings underscore the need for international consensus on relapse definitions and standardized relapse detection approaches to strengthen future clinical trials and optimize long-term disease management.

## DECLARATIONS

### Data availability statement

The authors confirm that the data supporting the findings of this study are available within the article and its supplementary materials.

### Ethical considerations

This research did not require ethical committee approval, as it is entirely based on previously published studies.

### Patient and Public Involvement statement

This study is a systematic review and meta-analysis based on previously published data, with no new primary data collected from patients. As such, patients and the public were not involved in the design, conduct, reporting, or dissemination plans of this research.

## Acknowledgements

We acknowledge the contributions of Professor Karan Madan, Department of Pulmonary, Critical Care and Sleep Medicine, All India Institute of Medical Sciences, New Delhi, India, and Professor Sahajal Dhooria, Department of Pulmonary Medicine, Postgraduate Institute of Medical Education and Research, Chandigarh, India, who kindly provided additional information regarding their respective studies in response to our inquiry. We also thank Mohammad Haider (MBBS Student, Autonomous State Medical College, Lakhimpur, Uttar Pradesh, India) for his assistance in drafting the tables and formatting the manuscript.

## AI declaration

During the preparation of this work the author(s) used Claude (Anthropic) in order to improve language and readability of the manuscript. After using this tool/service, the author(s) reviewed and edited the content as needed and take(s) full responsibility for the content of the published article.

## Author contributions

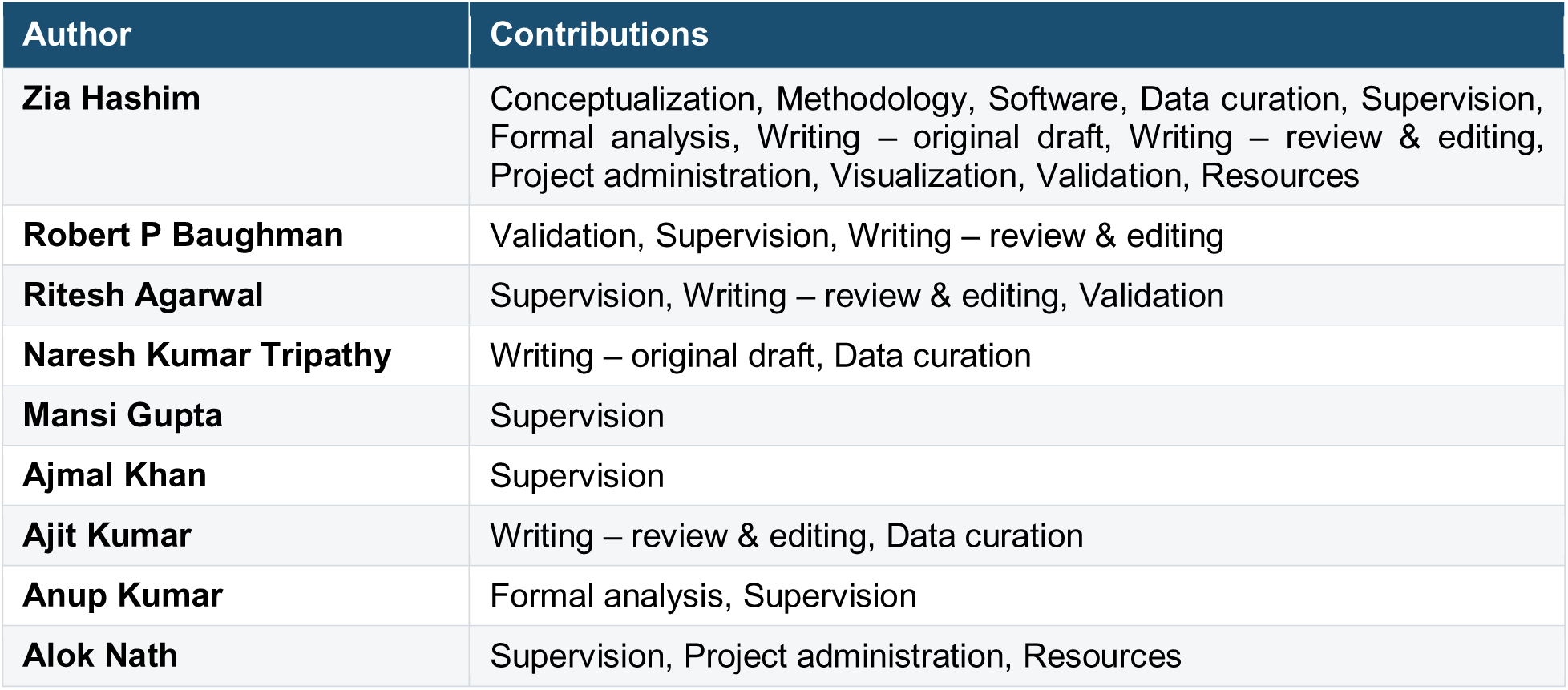

## SUPPLEMENTARY MATERIAL

## SUPPLEMENTARY TABLES

- Search Strategy
- Table S1: Freeman-Tukey Arcsine Transformation
- Table S2: Leave-One-Out Sensitivity Analysis
- Table S3: Galbraith Plot Data and Statistical Notes
- Table S4: Sarcoidosis Relapse Definitions
- **Table S5.** Timeframe Sensitivity Analysis - Impact of Changing the Early/Late Relapse Cutoff on Pooled Relapse Proportion
- Table S6: Meta-regression Analysis Results
- Table S7: GRADE Assessment

## SEARCH STRATEGY

A comprehensive systematic search was conducted across four major databases to identify relevant studies on pulmonary sarcoidosis relapse. Searches were conducted between January 26 and January 31, 2026. The search strategies for each database are detailed below:

**PubMed Search Strategy: 336 results on Jan 26, 2026**

~~~
((“Sarcoidosis, Pulmonary”[Mesh] OR “pulmonary sarcoidosis”[Title/Abstract] OR “lung sarcoidosis”[Title/Abstract] OR “sarcoidosis”[Title/Abstract]) AND (“Recurrence”[Mesh] OR “relapse”[Title/Abstract] OR “recurrence”[Title/Abstract] OR “flare-up”[Title/Abstract] OR “exacerbation”[Title/Abstract]) AND (“Prevalence”[Mesh] OR “prevalence”[Title/Abstract] OR “incidence”[Title/Abstract] OR “frequency”[Title/Abstract] OR “rate”[Title/Abstract] OR “Risk Factors”[Mesh] OR “risk factors”[Title/Abstract] OR “predictors”[Title/Abstract] OR “associations”[Title/Abstract] OR “determinants”[Title/Abstract]))
~~~

**Scopus Search Strategy: 345 results on Jan 26, 2026**

~~~
TITLE-ABS-KEY ((“pulmonary sarcoidosis” OR “lung sarcoidosis” OR “sarcoidosis”) AND (“relapse” OR “recurrence” OR “flareup” OR “exacerbation”) AND (“prevalence” OR “incidence” OR “frequency” OR “rate” OR “risk factors” OR “predictors” OR “associations” OR “determinants”)) OR INDEXTERMS (“Sarcoidosis, Pulmonary” OR “Recurrence” OR “Prevalence” OR “Risk Factors”)
~~~

**Embase Search Strategy: 730 results on Jan 26, 2026**

~~~
(’lung sarcoidosis’/exp OR ‘pulmonary sarcoidosis’:ti,ab,kw OR ‘lung sarcoidosis’:ti,ab,kw OR ‘sarcoidosis’:ti,ab,kw) AND (’disease recurrence’/exp OR ‘relapse’:ti,ab,kw OR ‘recurrence’:ti,ab,kw OR ‘flareup’:ti,ab,kw OR ‘exacerbation’:ti,ab,kw) AND (’prevalence’/exp OR ‘prevalence’:ti,ab,kw OR ‘incidence’:ti,ab,kw OR ‘frequency’:ti,ab,kw OR ‘rate’:ti,ab,kw OR ‘risk factor’/exp OR ‘risk factors’:ti,ab,kw OR ‘predictors’:ti,ab,kw OR ‘associations’:ti,ab,kw OR ‘determinants’:ti,ab,kw)
~~~

**Google Scholar Search: 549 results on Jan 26, 2026**

~~~
(“pulmonary sarcoidosis” OR “lung sarcoidosis” OR sarcoidosis) AND (relapse OR recurrence OR “flare up” OR “flare-up” OR flareup OR exacerbation) AND (prevalence OR incidence OR frequency OR rate OR “risk factors” OR predictors OR associations OR determinants)
~~~

**ClinicalTrials.gov Search Strategy: 6 records on January 31, 2026**

- Condition or disease: “sarcoidosis”
- Other terms: “relapse” OR “recurrence” OR “exacerbation” *(Advanced search at* https://clinicaltrials.gov/search)

**PROSPERO Search Strategy: 3 records on January 31, 2026**

- (“pulmonary sarcoidosis” OR “sarcoidosis”) AND (“relapse” OR “recurrence” OR “exacerbation”) *(Search at* https://www.crd.york.ac.uk/prospero)

**Gray Literature Search (ATS/ERS Conference Abstracts 2010–2025): 3 records on January 31, 2026**

- Manual screening of American Thoracic Society (ATS) and European Respiratory Society (ERS) annual conference abstract books (2010–2025) using the terms “sarcoidosis” AND (“relapse” OR “recurrence”)

**Table S1:**
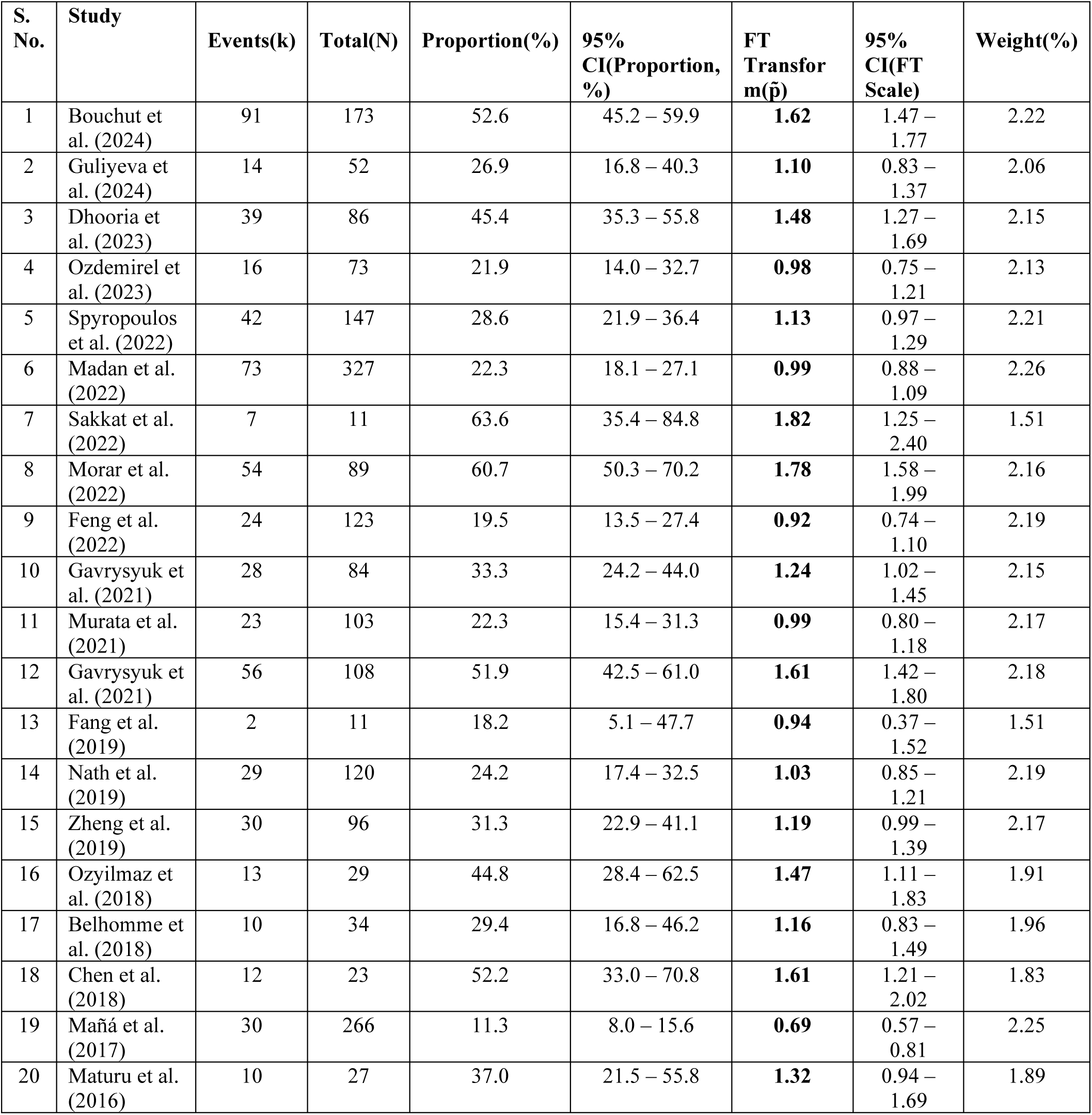
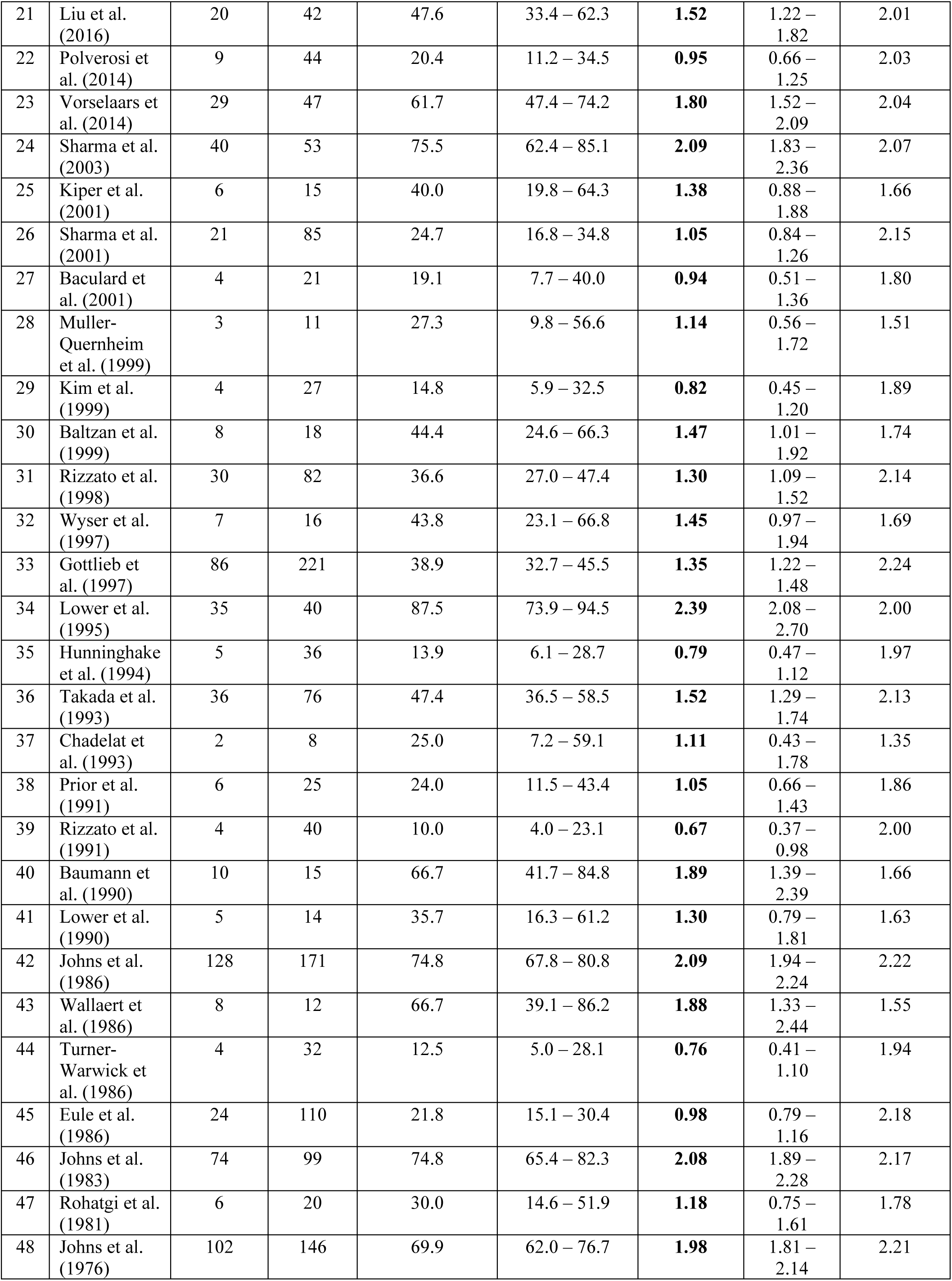
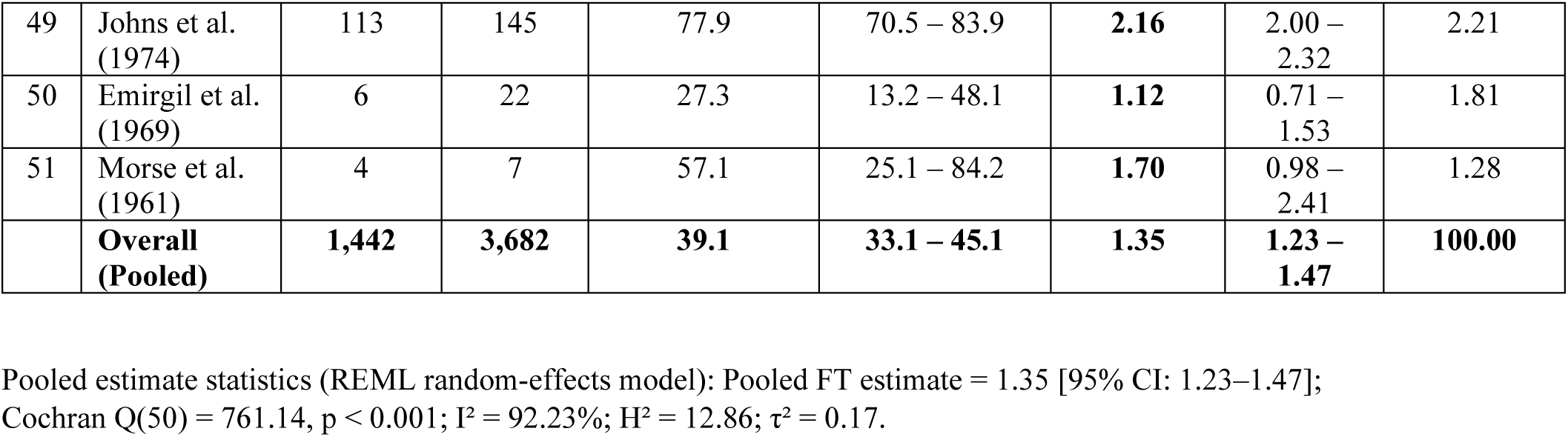
Freeman-Tukey Arcsine Transformation.

**Table S2:** Leave-One-Out Sensitivity Analysis.

| Omitted Study No. | Pooled Prevalence | Lower 95% CI | Upper 95% CI |
| --- | --- | --- | --- |
| 50 | 0.397 | 0.330 | 0.464 |
| 49 | 0.386 | 0.323 | 0.449 |
| 48 | 0.388 | 0.323 | 0.454 |
| 47 | 0.397 | 0.330 | 0.464 |
| 46 | 0.387 | 0.322 | 0.452 |
| 45 | 0.399 | 0.331 | 0.466 |
| 44 | 0.401 | 0.334 | 0.468 |
| 43 | 0.390 | 0.324 | 0.457 |
| 42 | 0.387 | 0.323 | 0.451 |
| 41 | 0.396 | 0.329 | 0.462 |
| 40 | 0.390 | 0.324 | 0.457 |
| 39 | 0.401 | 0.335 | 0.468 |
| 38 | 0.398 | 0.331 | 0.465 |
| 37 | 0.395 | 0.329 | 0.461 |
| 36 | 0.393 | 0.326 | 0.461 |
| 35 | 0.400 | 0.334 | 0.467 |
| 34 | 0.384 | 0.320 | 0.449 |
| 33 | 0.395 | 0.327 | 0.464 |
| 32 | 0.394 | 0.327 | 0.461 |
| 31 | 0.396 | 0.328 | 0.463 |
| 30 | 0.394 | 0.327 | 0.461 |
| 29 | 0.400 | 0.333 | 0.467 |
| 28 | 0.397 | 0.330 | 0.464 |
| 27 | 0.399 | 0.332 | 0.466 |
| 26 | 0.398 | 0.331 | 0.466 |
| 25 | 0.395 | 0.328 | 0.462 |
| 24 | 0.387 | 0.321 | 0.453 |
| 23 | 0.390 | 0.324 | 0.457 |
| 22 | 0.399 | 0.332 | 0.466 |
| 21 | 0.393 | 0.326 | 0.460 |
| 20 | 0.395 | 0.328 | 0.462 |
| 19 | 0.401 | 0.337 | 0.466 |
| 18 | 0.393 | 0.326 | 0.459 |
| 17 | 0.397 | 0.330 | 0.464 |
| 16 | 0.394 | 0.327 | 0.461 |
| 15 | 0.397 | 0.329 | 0.464 |
| 14 | 0.398 | 0.331 | 0.466 |
| 13 | 0.399 | 0.332 | 0.466 |
| 12 | 0.392 | 0.325 | 0.460 |
| 11 | 0.399 | 0.331 | 0.466 |
| **10** | 0.396 | 0.329 | 0.464 |
| **9** | 0.399 | 0.332 | 0.467 |
| **8** | 0.390 | 0.324 | 0.457 |
| **7** | 0.391 | 0.324 | 0.458 |
| **6** | 0.399 | 0.330 | 0.468 |
| **5** | 0.397 | 0.330 | 0.465 |
| **4** | 0.399 | 0.332 | 0.466 |
| **3** | 0.394 | 0.326 | 0.461 |
| **2** | 0.398 | 0.330 | 0.465 |
| **1** | 0.392 | 0.325 | 0.460 |

**Table S3:**
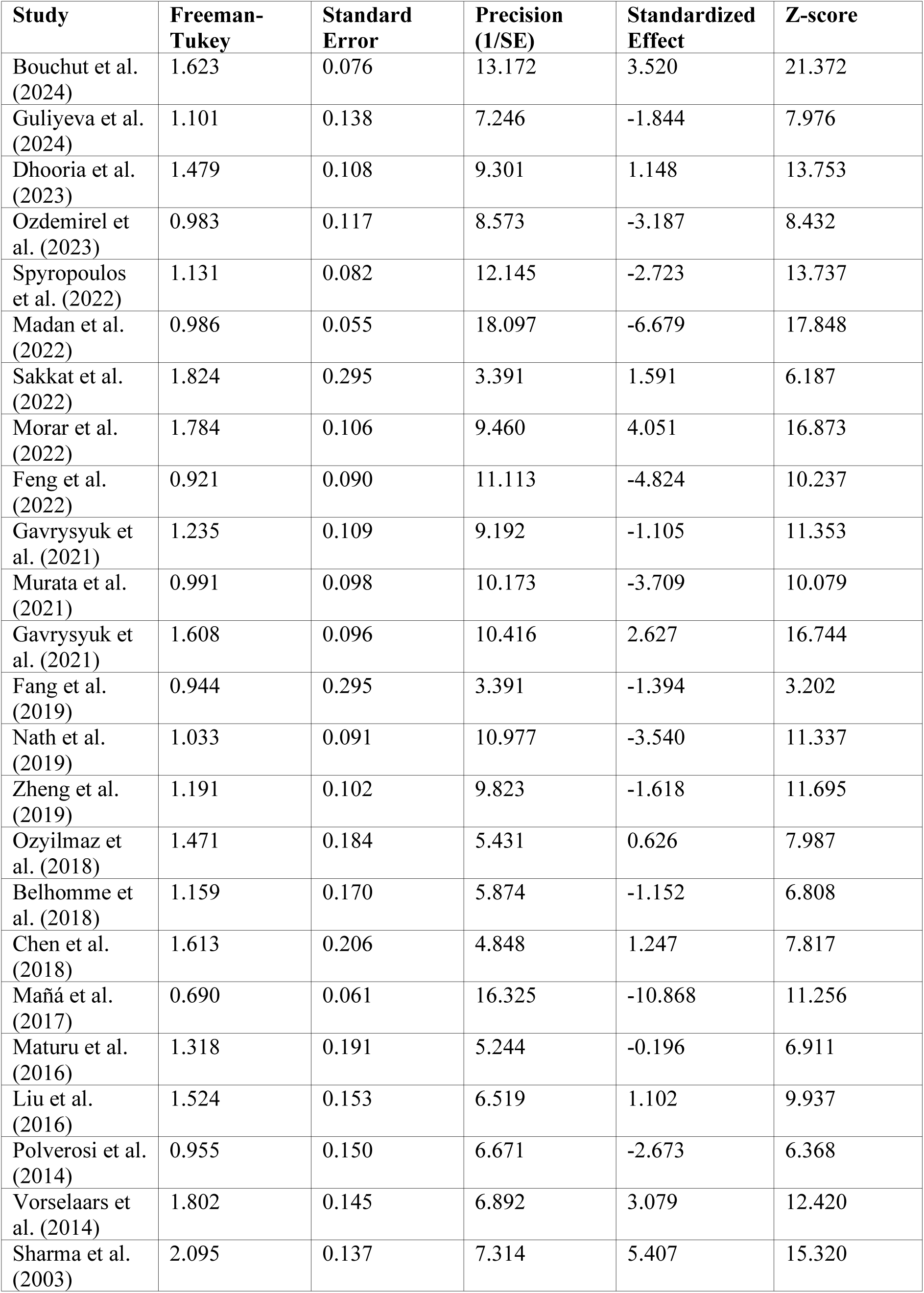
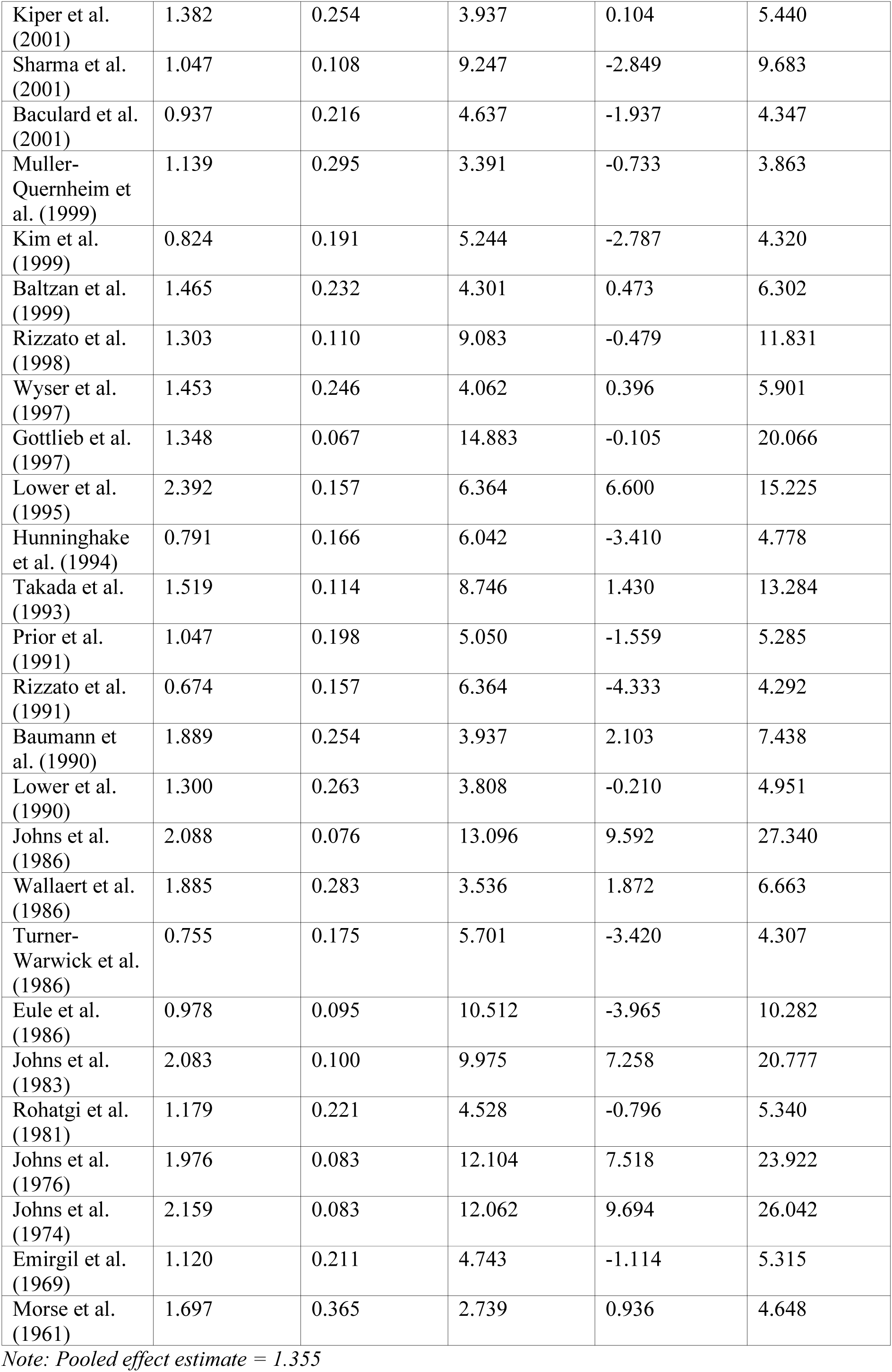
Galbraith Plot Data. Data for constructing Galbraith plot to assess heterogeneity in the meta-analysis. Precision = 1/SE; Standardized effect = (Effect - Pooled effect)/SE.

**Table S4.**
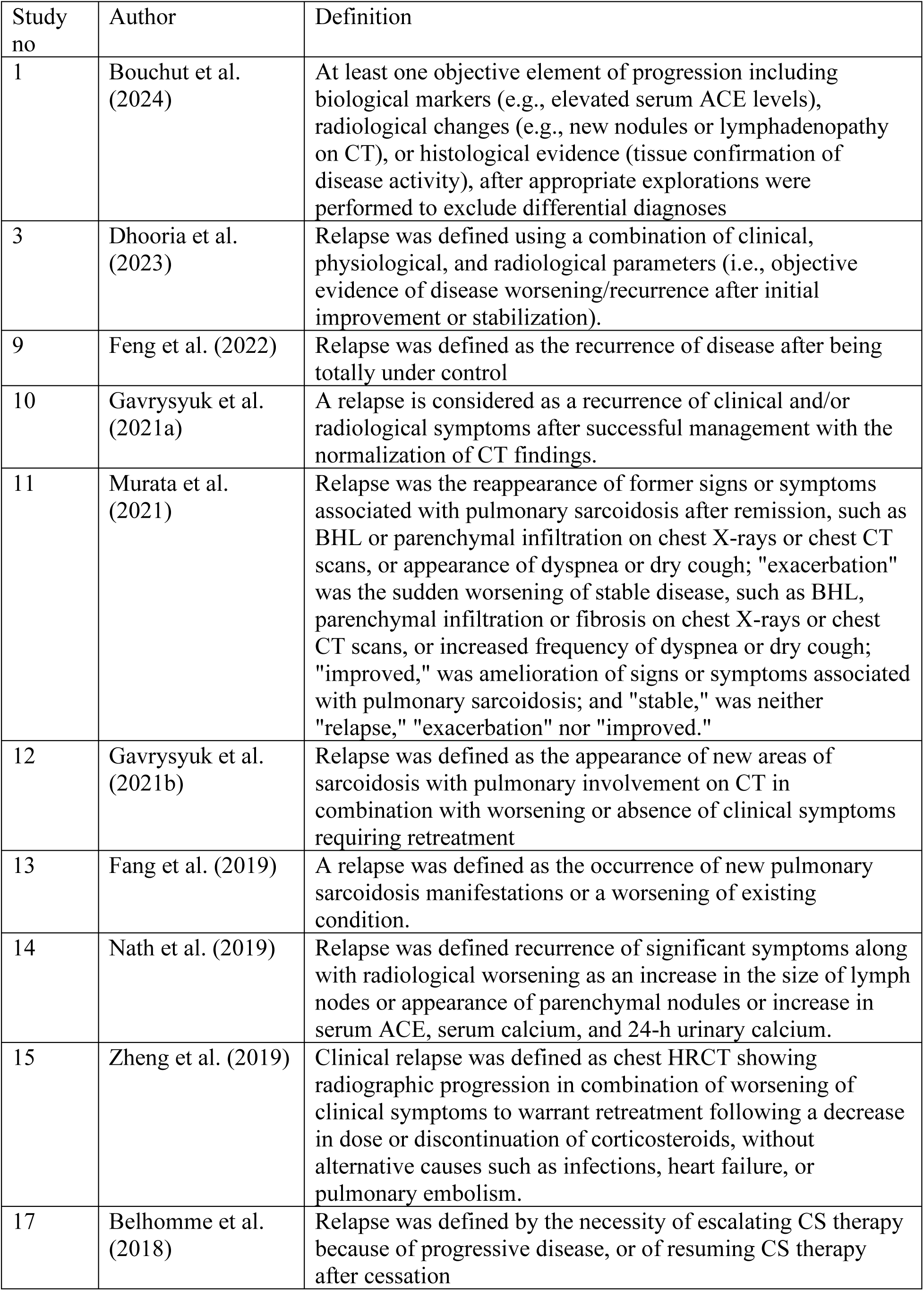
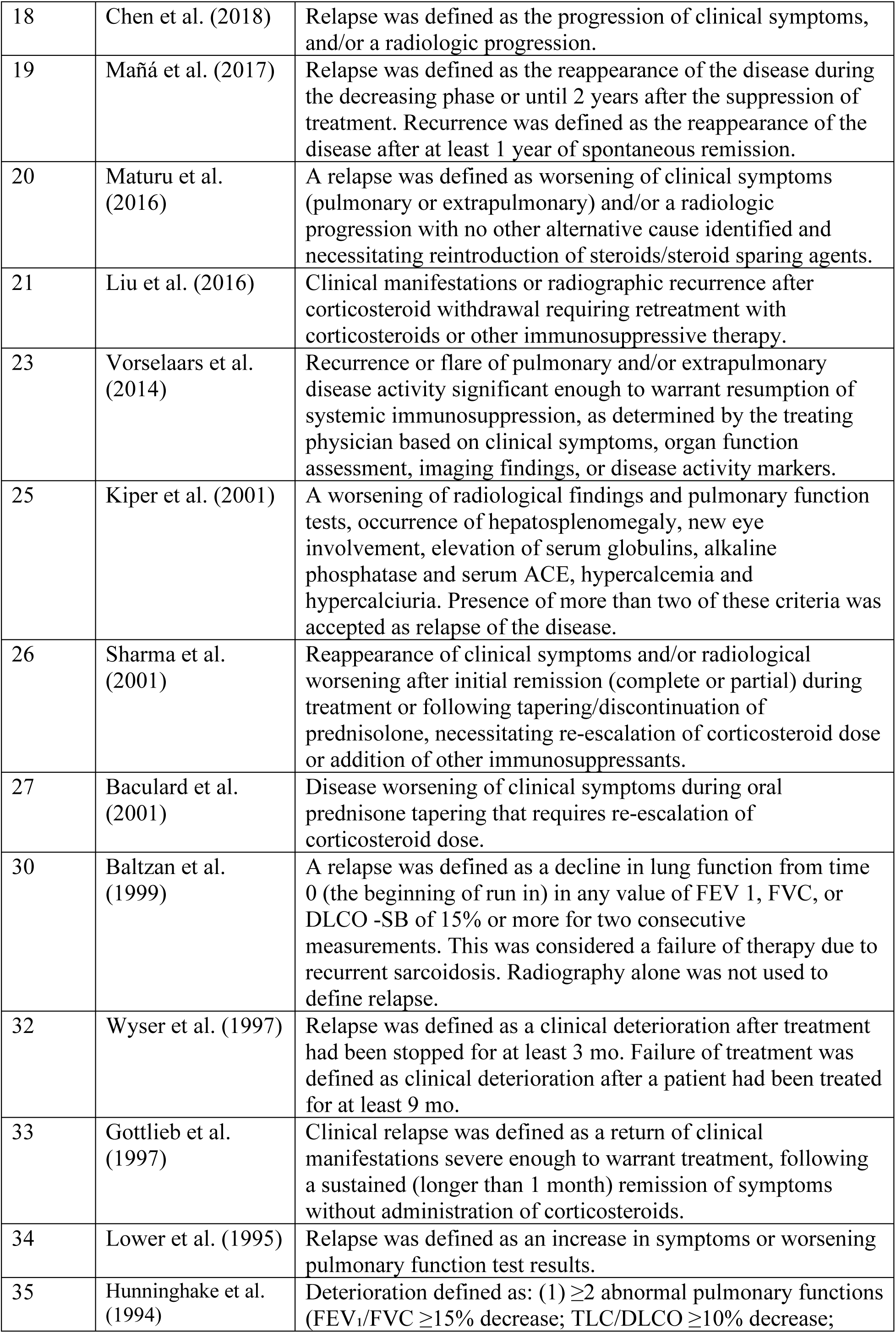
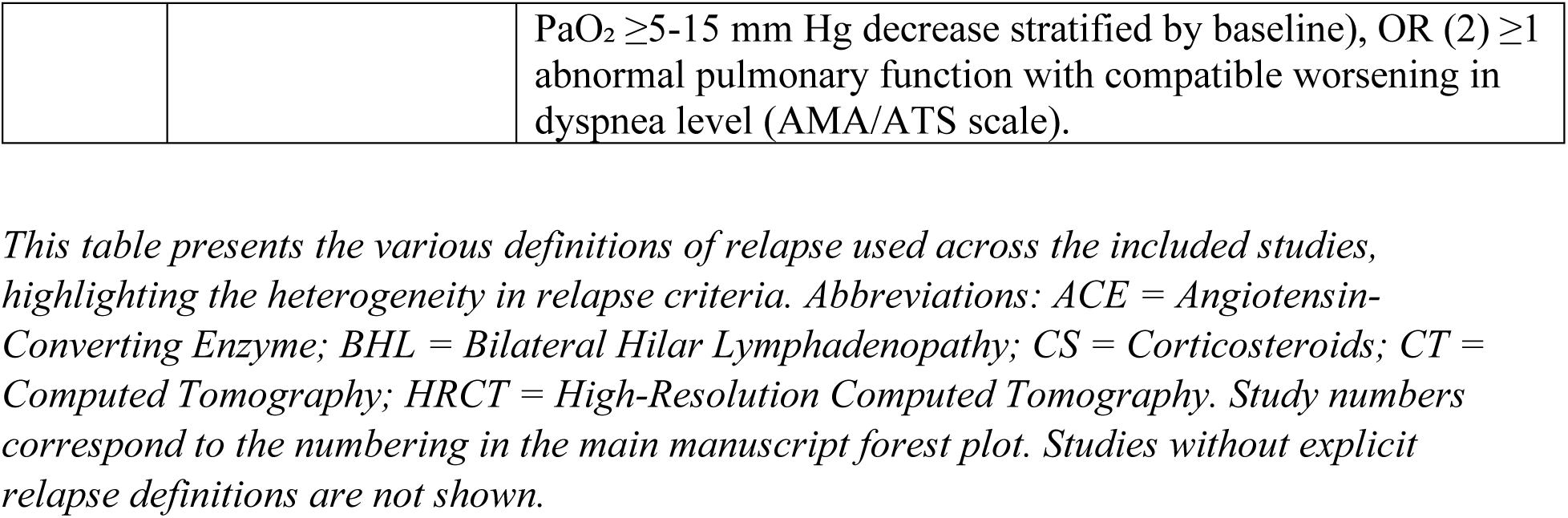
Definitions of Pulmonary Sarcoidosis Relapse Used in Included Studies.

**Table S5.** Timeframe Sensitivity Analysis - Impact of Changing the Early/Late Relapse Cutoff on Pooled Relapse Proportion.

| Cutoff | Studies (n) | Pooled Proportion | 95% CI | I<sup>2</sup> (%) |
| --- | --- | --- | --- | --- |
| ≥1 month (Primary) | 19 | 0.446 | 0.370–0.525 | 84.4% |
| ≥2 months | 15 | 0.431 | 0.367–0.497 | 72.3% |
| ≥3 months | 11 | 0.469 | 0.405–0.534 | 54.9% |
| ≥6 months | 8 | 0.468 | 0.395–0.542 | 63.6% |

*Analysis restricted to the 19 studies that reported numeric time-to-relapse ranges. Primary all-51-study pooled estimate: 0.39 (95% CI 0.33–0.45). I<sup>2</sup> = I-squared statistic for heterogeneity. CI = confidence interval. Pooled proportions estimated using the Random-effects model*

**Table S6.** Meta-regression Analysis Results. *Meta-regression was performed to identify potential sources of heterogeneity. Study design was the only significant factor identified*.

| Variable | Coefficient (95% CI) | Standard Error | z | p-value |
| --- | --- | --- | --- | --- |
| **Multivariable meta-regression (N = 51 studies; REML random-effects)** |  |  |  |  |
| Relapse definition (explicit vs. absent) | 0.010<br>(−0.139 to 0.160) | 0.076 | 0.14 | 0.892 |
| Publication era (pre-2000 vs. post-2000) | 0.056<br>(−0.100 to 0.212) | 0.079 | 0.71 | 0.480 |
| Study design (retrospective vs. prospective) | 0.148<br>(0.031 to 0.265) | 0.060 | 2.47 | **0.014** |
| Geographic region (non-Asian vs. Asian) | 0.086<br>(−0.059 to 0.232) | 0.074 | 1.16 | 0.245 |
| Sample size (>50 vs. ≤50 patients) | −0.043<br>(−0.172 to 0.086) | 0.066 | −0.65 | 0.515 |
| Study quality (moderate vs. low risk of bias) | −0.051<br>(−0.213 to 0.110) | 0.082 | −0.62 | 0.533 |

| Univariate meta-regression: treatment duration as sole predictor (N = 30 studies†) |  |  |  |  |
| --- | --- | --- | --- | --- |
| Treatment duration†<br>(>18 vs. ≤18 months) | -0.086<br>(-0.183 to 0.012) | 0.050 | -1.72 | 0.085 |

Abbreviations: CI, confidence interval; REML, restricted maximum likelihood; SE, standard error.
† Treatment duration was not included as a covariate in the primary multivariable model because duration data were unavailable for 21 of 51 studies (41%). A separate pre-specified univariate meta-regression was performed, restricted to the 30 studies with estimable duration data, to evaluate the independent association of treatment duration with relapse prevalence.

**Table S7.** GRADE Assessment of Evidence Certainty. *The Grading of Recommendations Assessment, Development and Evaluation (GRADE) approach was used to assess the certainty of evidence*.

| Outcome | Studies (N) | Certainty | Reasons for Rating |
| --- | --- | --- | --- |
| Relapse Prevalence | 51 studies (3,682 followed up) | Low | Downgraded for bias and inconsistency |
| Relapse Prevalence (Prospective Studies) | 22 studies (1,737) | Moderate | Downgraded for bias and inconsistency |
| Risk by Black Race | Subset with data | Moderate | Downgraded for bias; upgraded for large effect |
| Association with Age | 15 studies (1,434) | Moderate | Downgraded for bias |
| Female Sex (vs Male) | 15 studies (1,326) | Moderate | Downgraded for bias |

## SUPPLEMENTARY FIGURES

**Quality Assessment and Heterogeneity Analysis**

- **Figure S1:** Hoy et al. Risk of Bias Assessment
- **Figure S2:** Study bias and effect on the relapse prevalence
- **Figure S3:** Sensitivity analysis across subgroups (design, relapse definition, quality, sample size, publication year, geographical location)
- **Figure S4:** Leave-one-out analysis
- **Figure S5:** Freeman-Tukey arcsine transformation
- **Figure S6:** Freeman-Tukey back transformation
- **Figure S7:** Baujat plot identifying influential studies
- **Figure S8:** Galbraith plot for heterogeneity assessment
- **Figure S9:** Excluding outliers - combined analysis

**Clinical Characteristics and Outcomes**

- **Figure S10:** Median time to relapse after corticosteroid cessation
- **Figure S11:** Pooled mean differences in age between relapsed and non-relapsed patients
- **Figure S12:** Log odds ratios for sex (male vs. female)
- **Figure S13:** Relapse rates between Black and White patients
- **Figure S14:** Relapse prevalence in adult-only populations
- **Figure S15:** Relapse rates across radiographic stages
- **Figure S16:** Relapse prevalence stratified by disease extent (isolated pulmonary vs. multiorgan)
- **Figure S17:** Relapse prevalence stratified by treatment type (corticosteroids, steroid-sparing agents, mixed)
- **Figure S18:** Subgroup analysis of relapse proportions in pulmonary sarcoidosis stratified by duration of corticosteroid therapy
- **Figure S19:** Effect of relapse definition criteria (clinical alone vs. clinical + additional criteria)

*Note: All forest plots display individual study estimates (squares sized by weight) with 95% confidence intervals (horizontal lines) and pooled estimates (diamonds) using random-effects REML model.*

**Figure S1:**
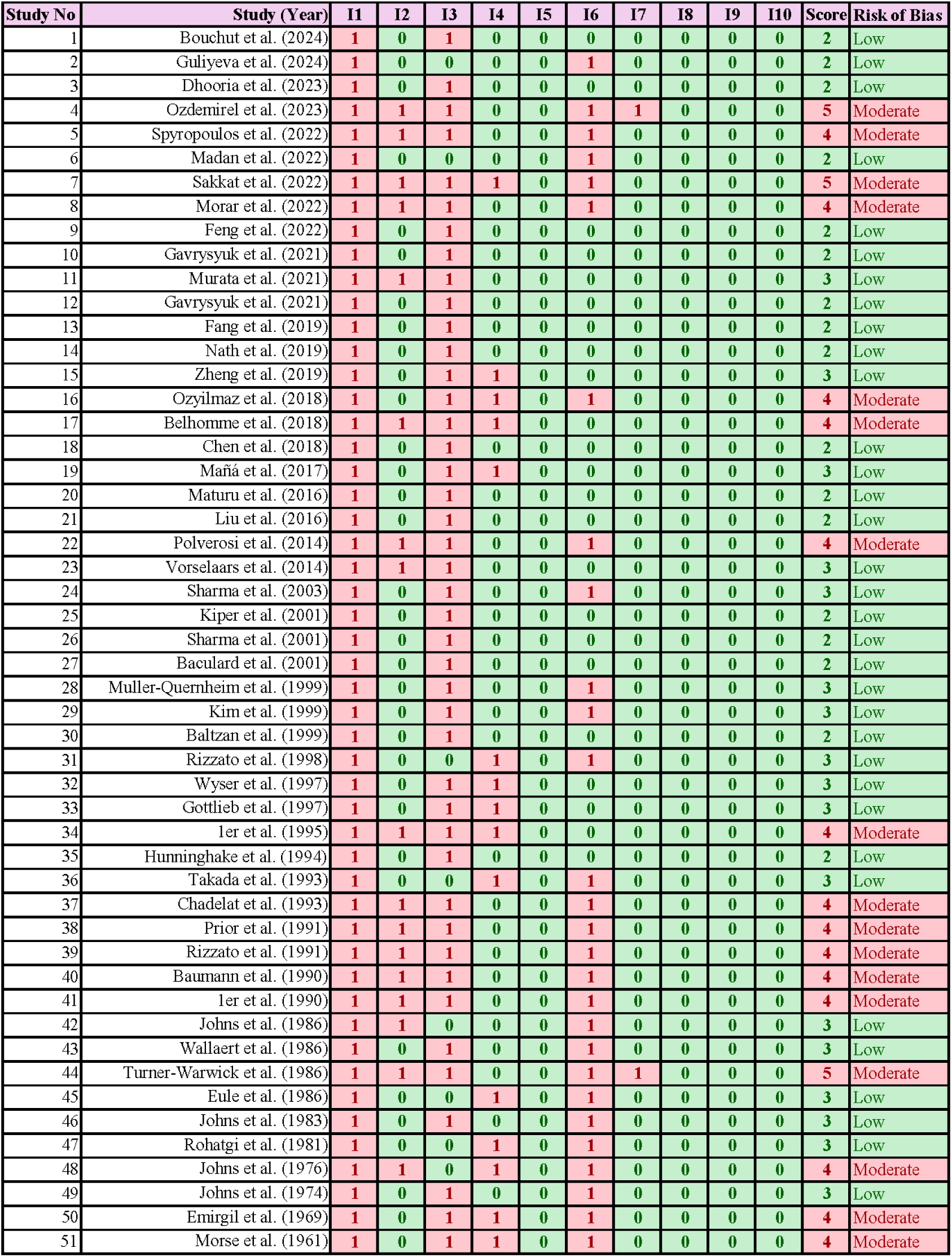
Hoy et al. Risk of Bias Assessment.

**Figure S2:**
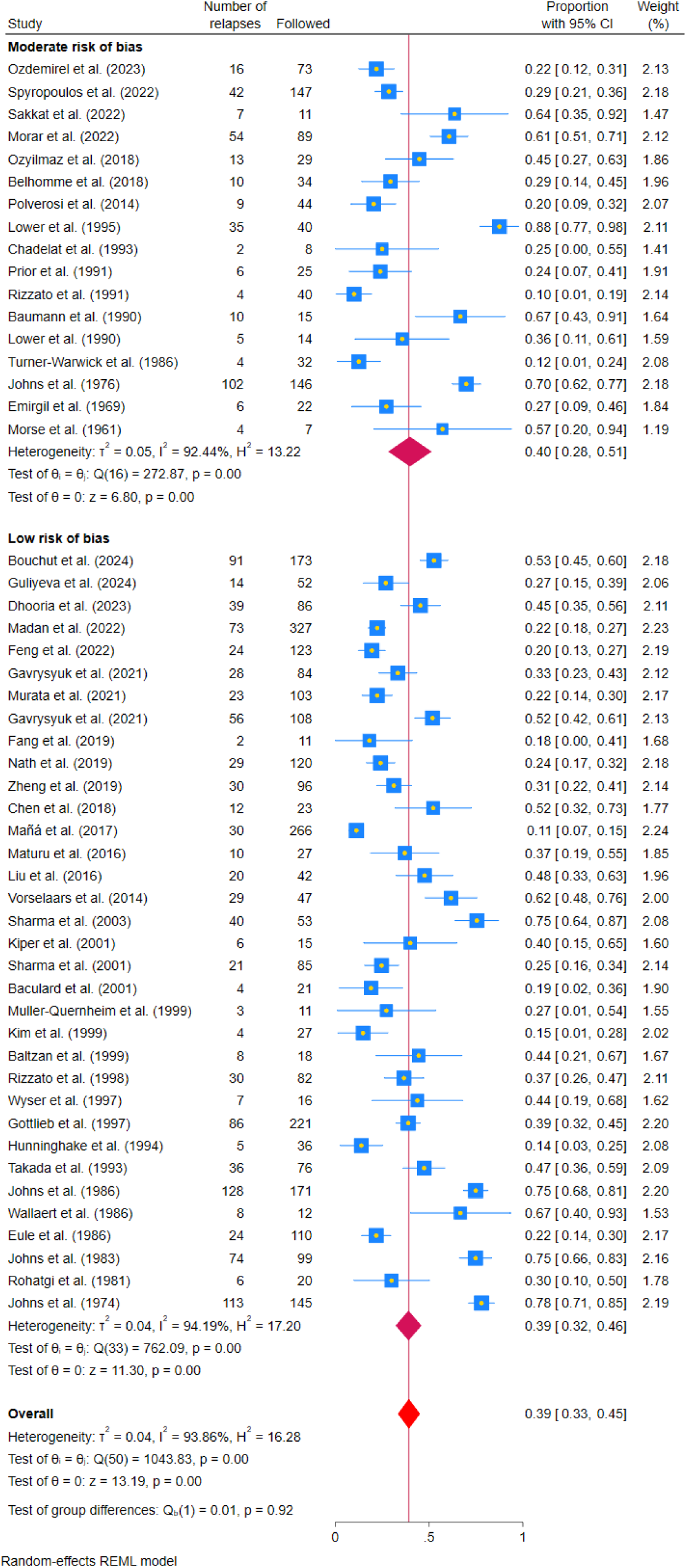
Study bias and effect on the relapse prevalence.

**Figure S3:**
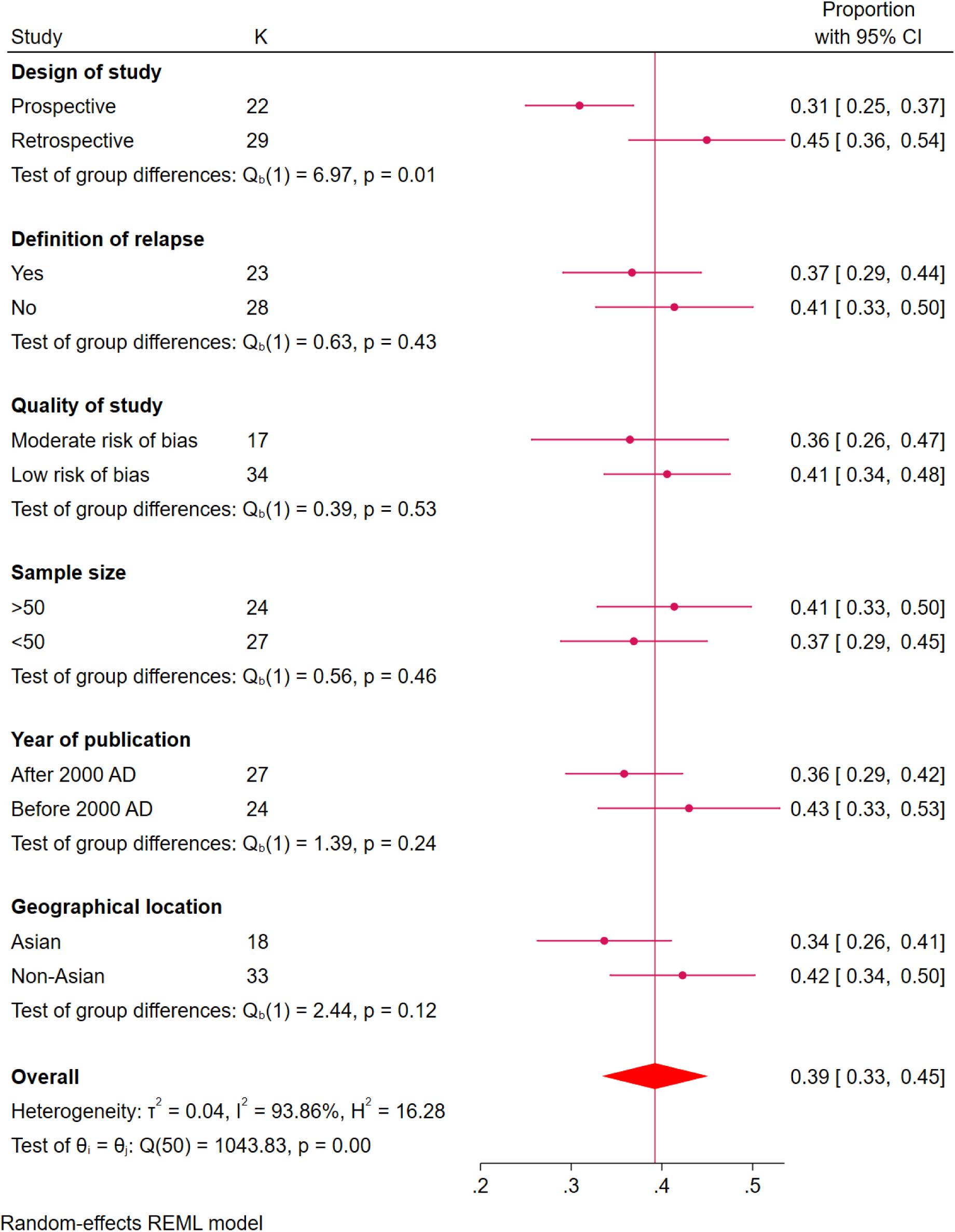
Sensitivity analysis: Forest plot showing the proportion of studies with 95% Cis across subgroups: design (prospective vs. retrospective), relapse definition (yes vs. no), year (after vs. before 2000), Asian vs. non-Asian, patients >50 (yes vs. no), and quality (low vs. moderate risk of bias).

**Figure S4:**
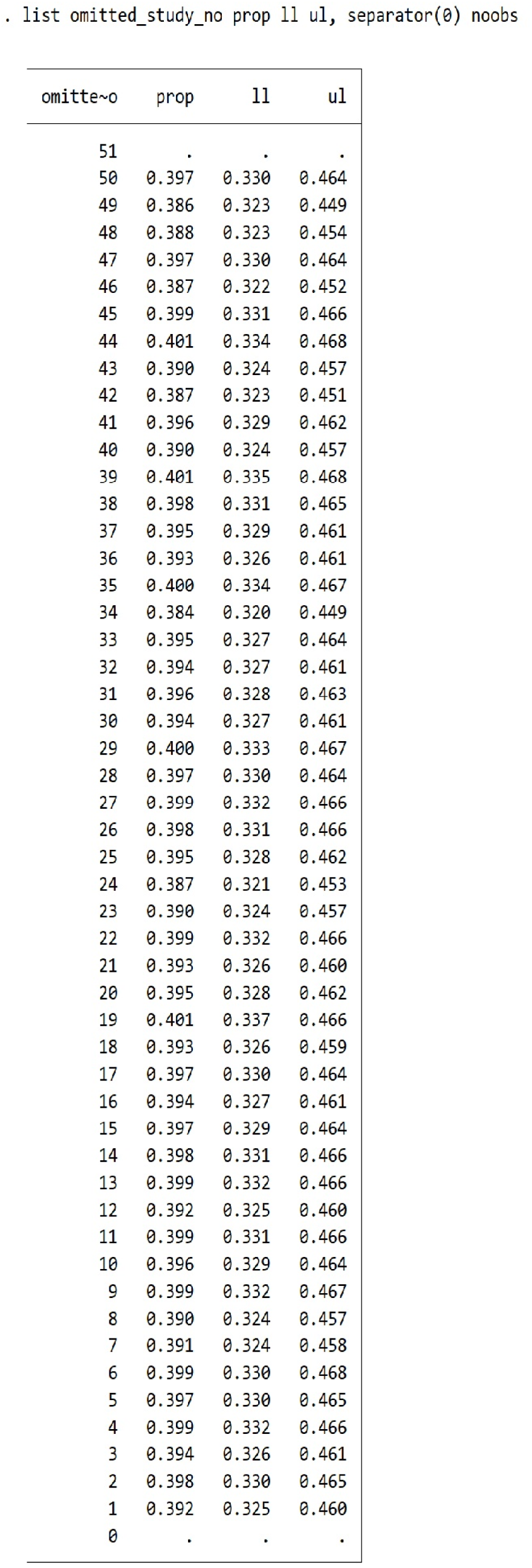
Leave-one-out analysis by sequentially excluding each study and recalculating the pooled prevalence. This analysis demonstrates the stability of the pooled estimate, with no single study having disproportionate influence on the overall results.

**Figure S5:**
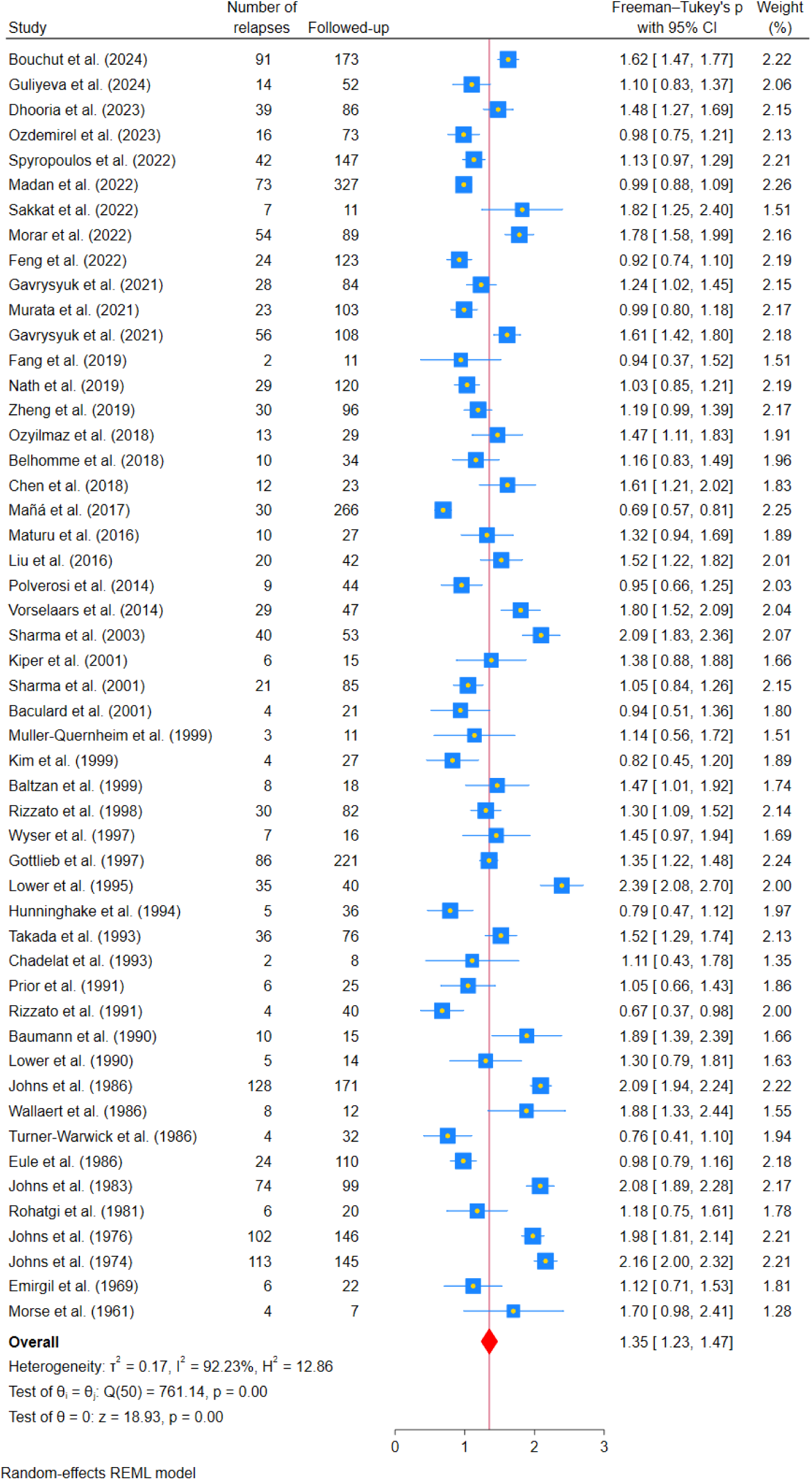
Freeman Tukey.

**Figure S6:**
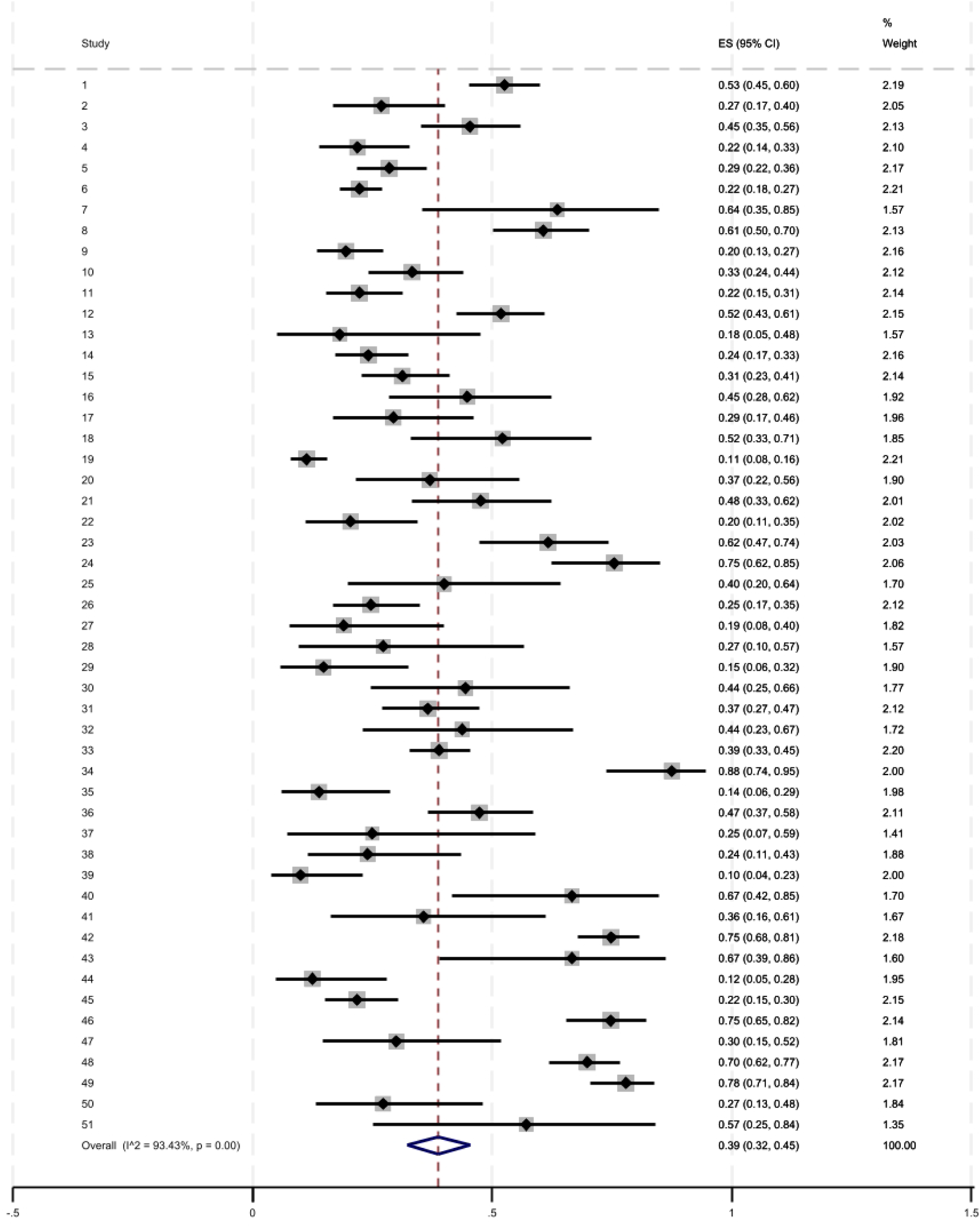
Freeman Tukey Back Transformation.

**Figure S7:**
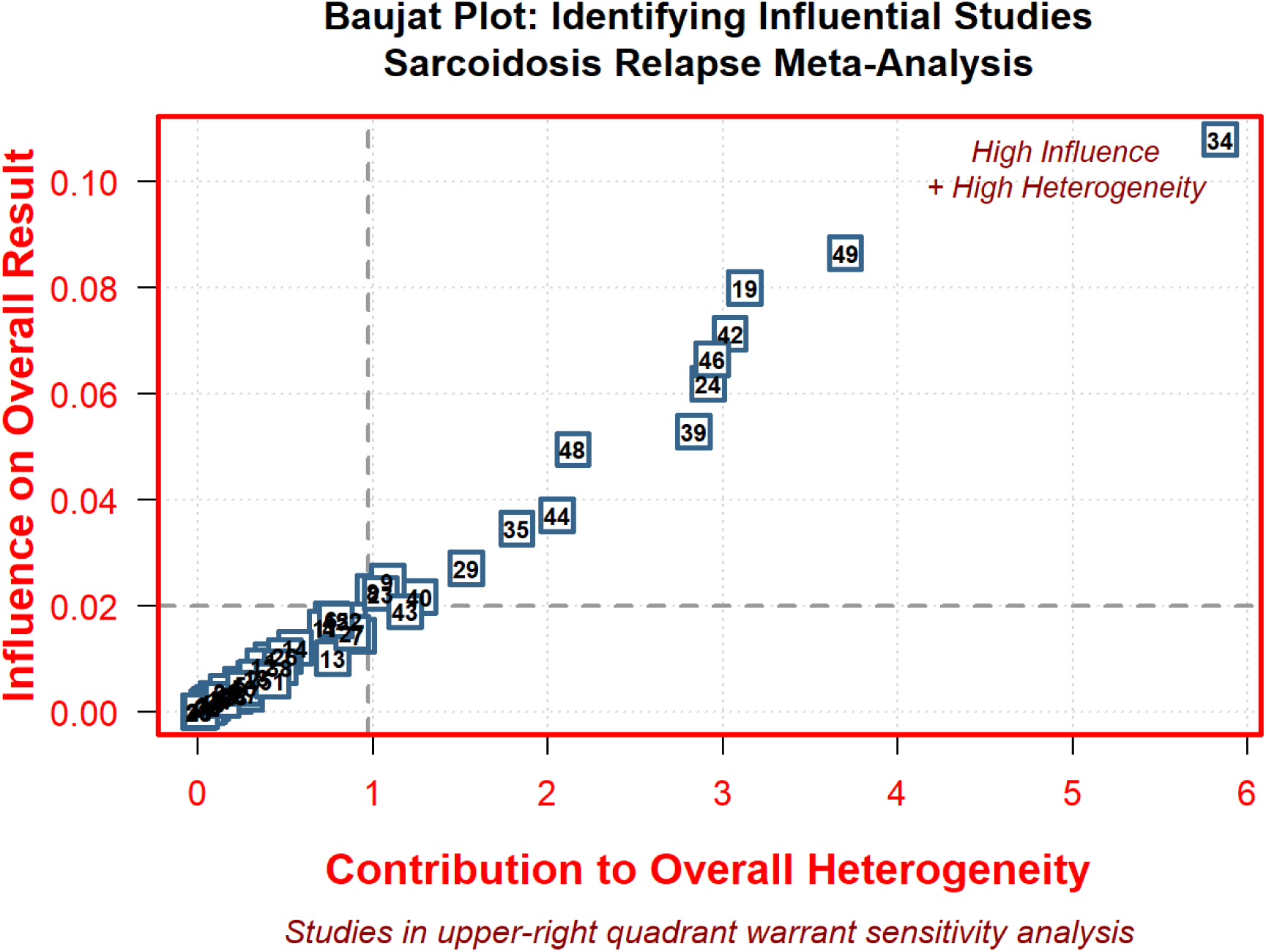
Baujat Plot Identifying Influential Studies in Sarcoidosis Relapse. Meta-Analysis, Studies in the upper-right quadrant contribute substantially to both overall heterogeneity and the pooled estimate. The following studies were identified as outliers warranting sensitivity analysis: Mañá et al. (2017, #19), Sharma et al. (2003, #24), Lower et al. (1995, #34), and Hunninghake et al. (1994, #35), Rizzato et al. (1991, #39), Johns et al. (1986, #42), Turner-Warwick et al. (1986, #44), Johns et al. (1983, #46), Johns et al. (1976, #48), and Johns et al. (1974, #49).

**Figure S8:**
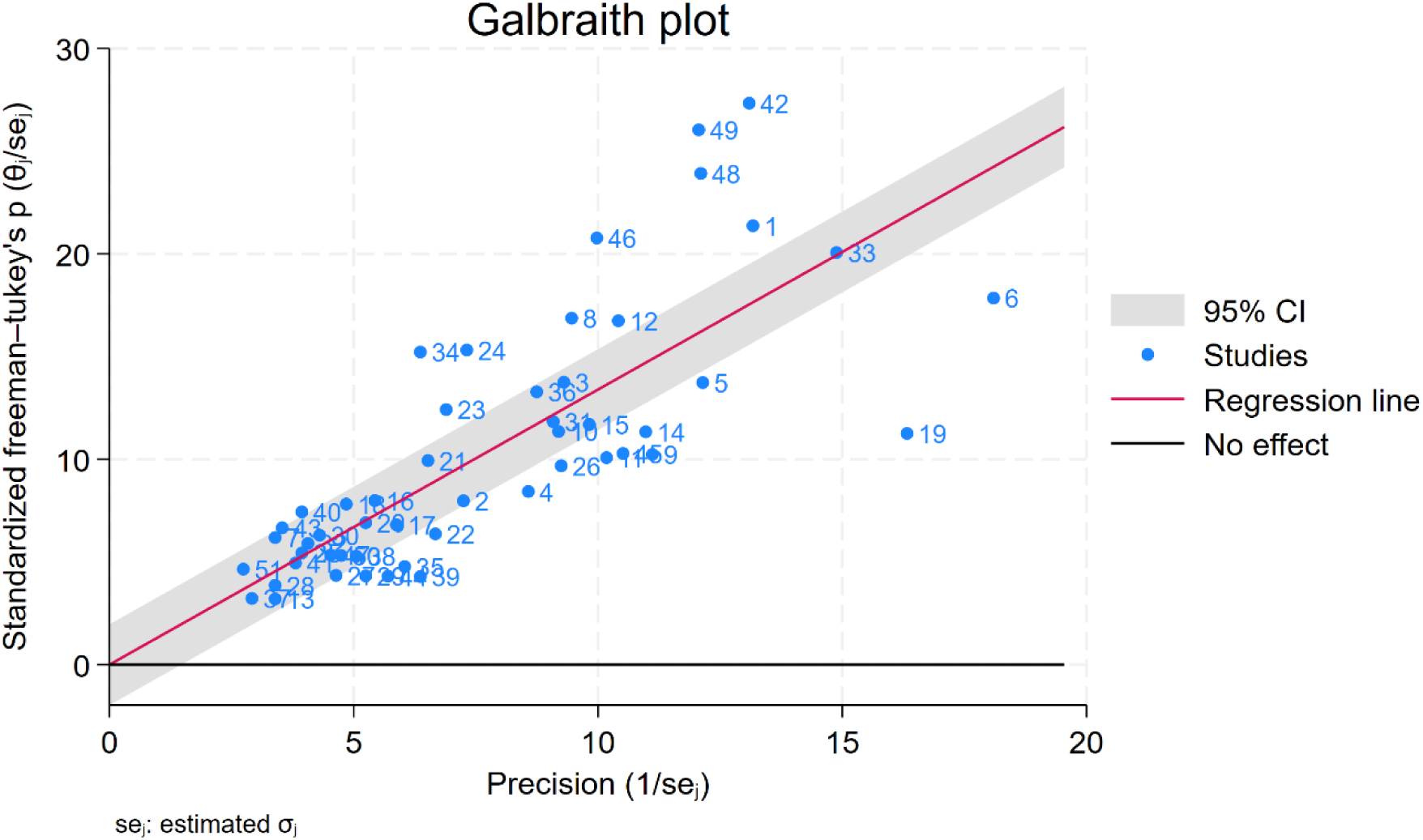
Galbraith plot shows the relationship between study precision (1/SE) and standardized effect size (θ/SE %), with labelled studies, a regression line, a 95% CI band, and a no-effect line to assess heterogeneity and identify outliers.

**Figure S9:**
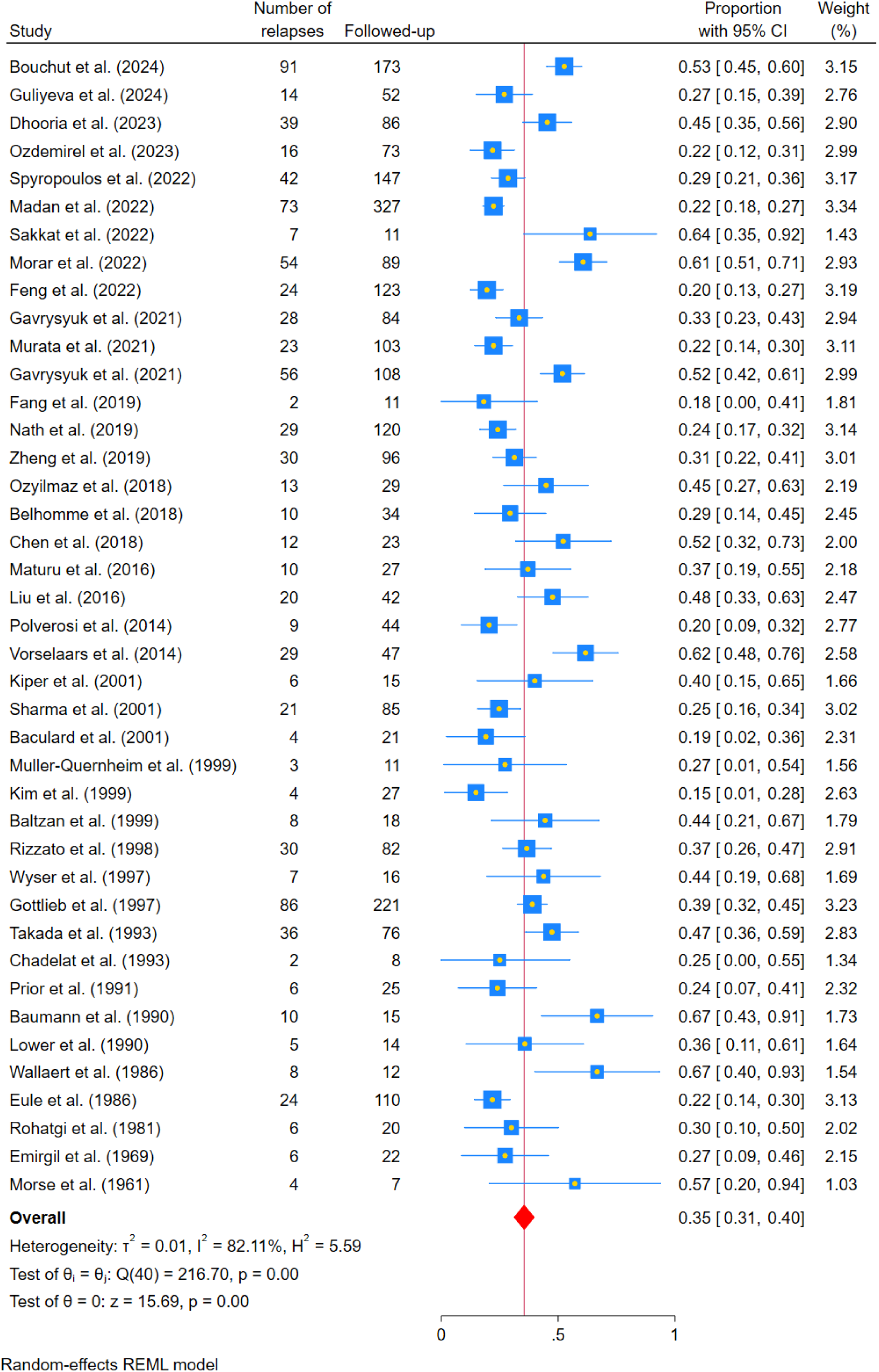
Excluding outliers from Baujat and Galbraith plots. The exclusion of studies 19, 24, 34, 35, 39, 42, 44, 46, 48, and 49 resulted in 82% heterogeneity; however, the pooled estimate demonstrated stability.

**Figure S10:**
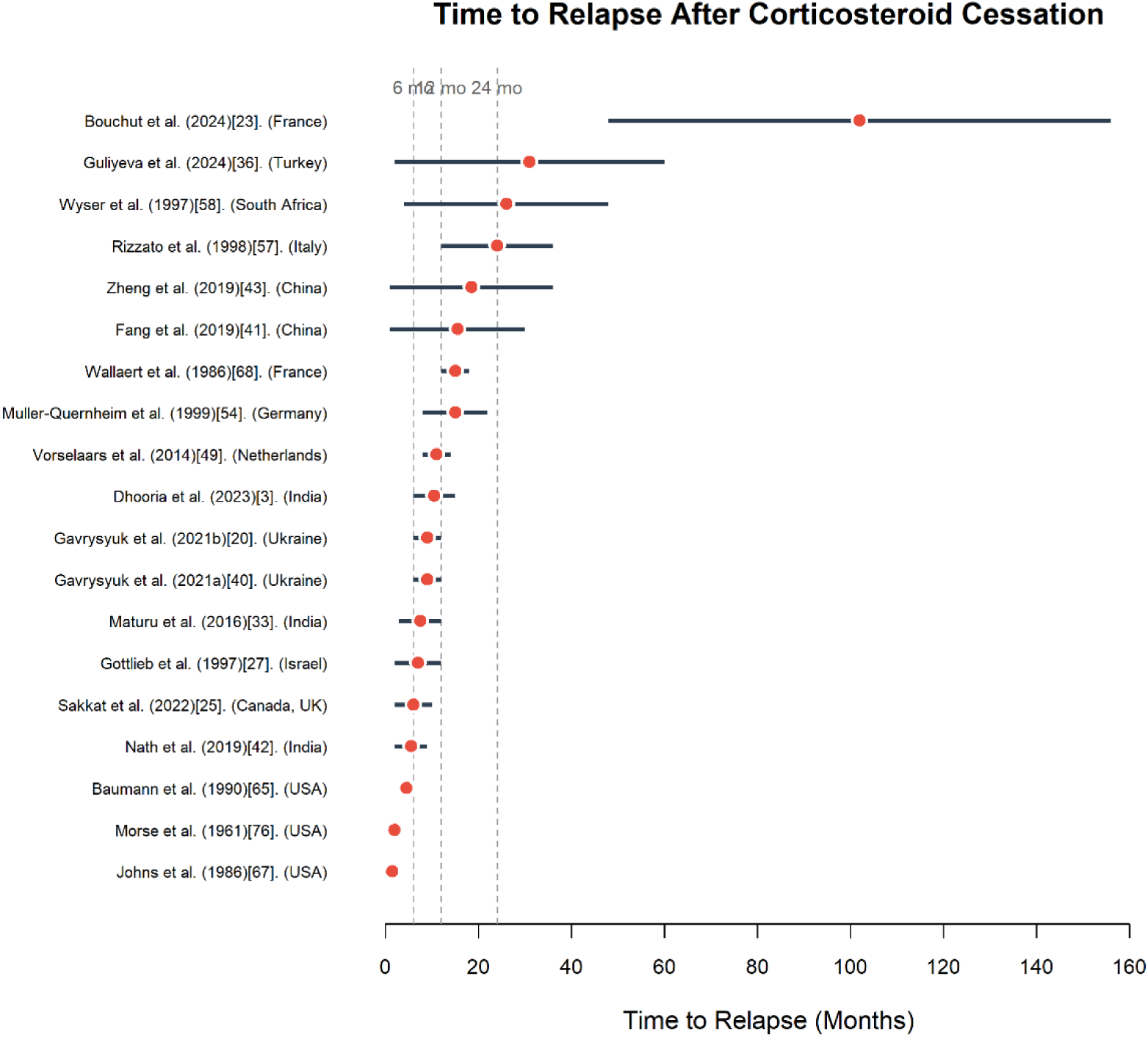
Forest plot showing median or mean time to relapse after corticosteroid cessation across 19 studies. Orange circles represent point estimates; horizontal lines represent confidence intervals. Studies are listed with first author, publication year, sample size in brackets, and country. Vertical dashed lines indicate reference points at 6 and 24 months. Time to relapse ranges from <10 months (Johns et al., Morse et al., Baumann et al.) to ∼100 months (Bouchut et al.). Studies arranged chronologically from oldest (bottom) to most recent (top).

**Figure S11:**
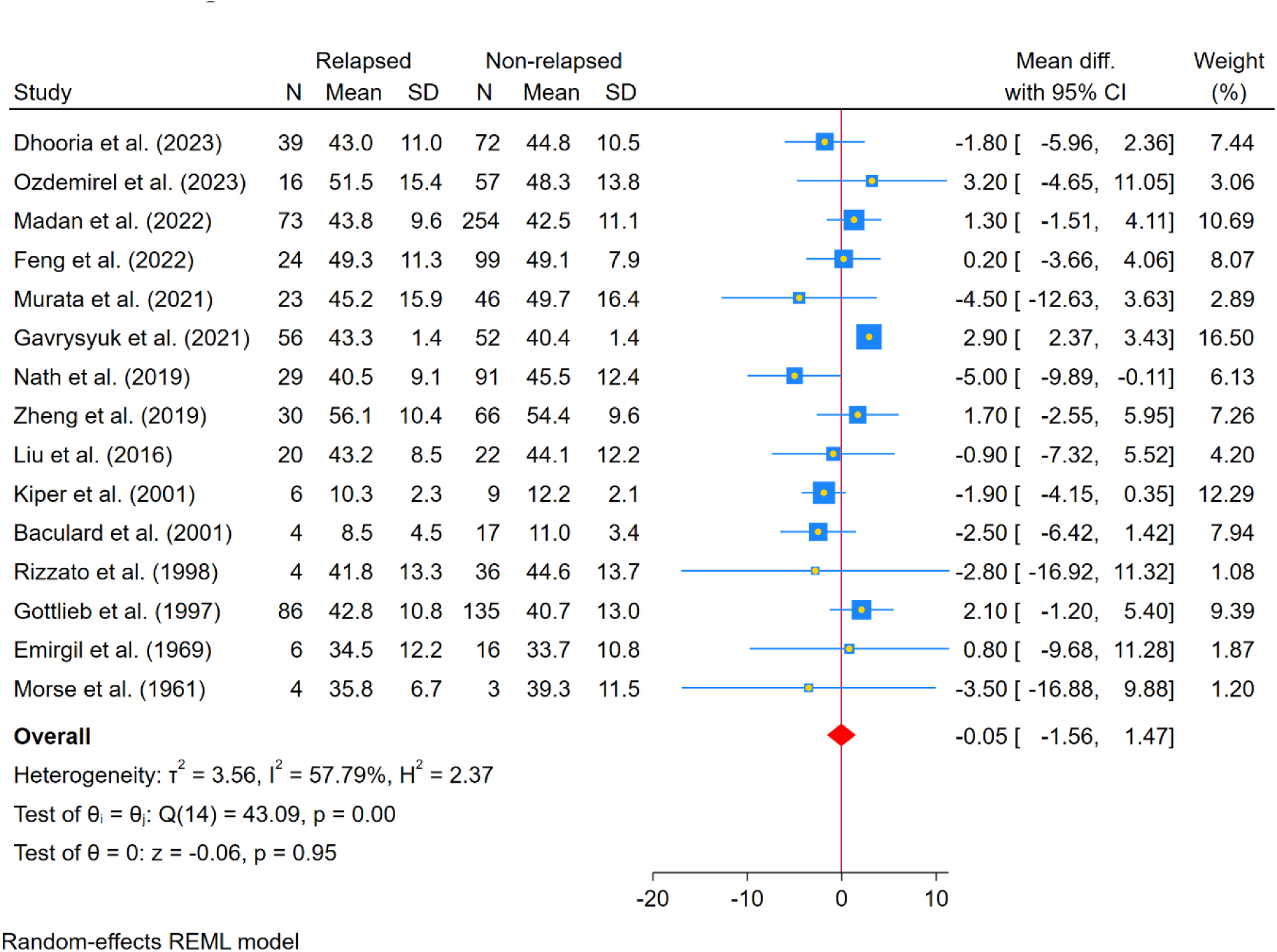
Forest plot showing pooled mean differences in the ages of patients with relapse versus non-relapse.

**Figure S12:**
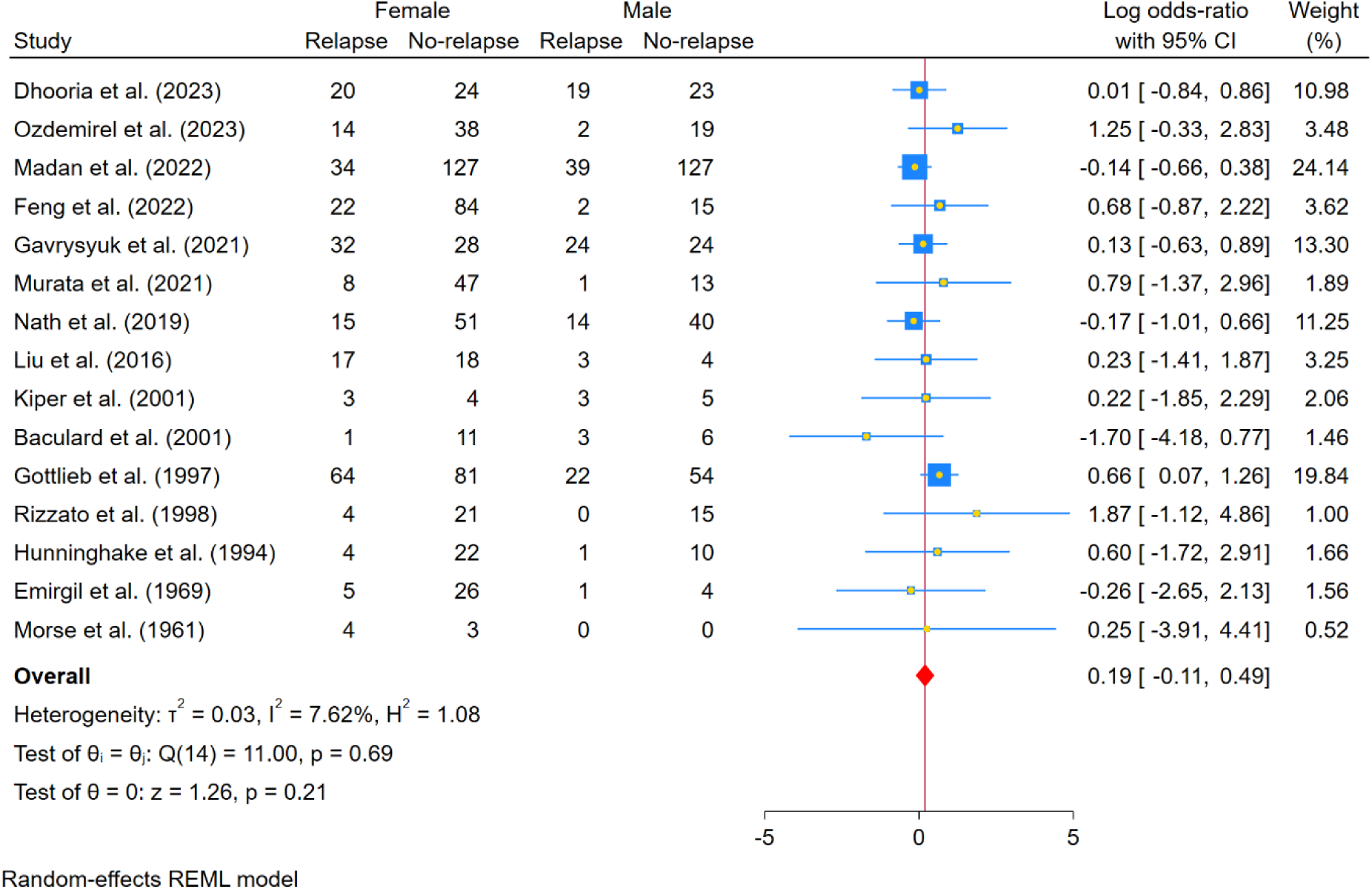
Log odds ratios for the sex of patients with pulmonary sarcoidosis.

**Figure S13:**
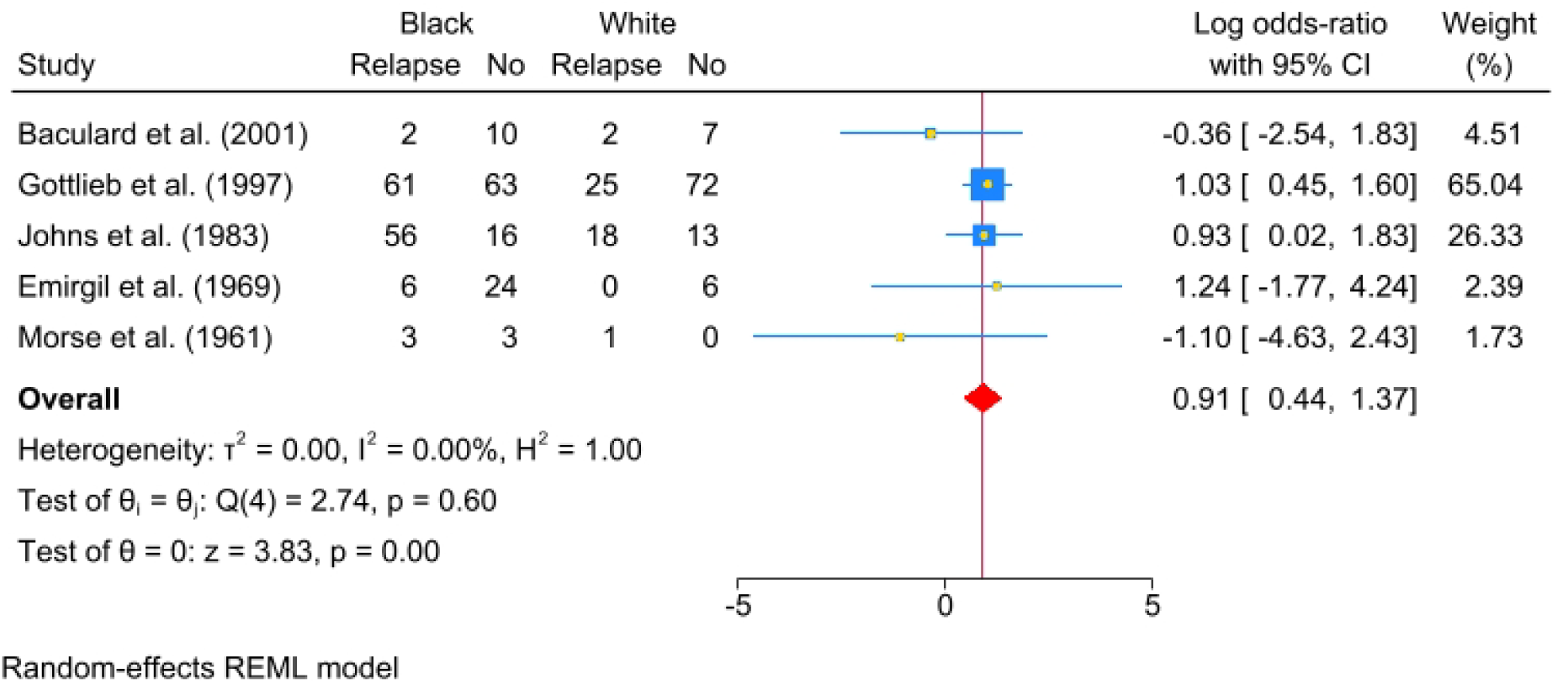
Forest plot comparing relapse rates between Black and White patients with pulmonary sarcoidosis (Random-effects model).

**Figure S14:**
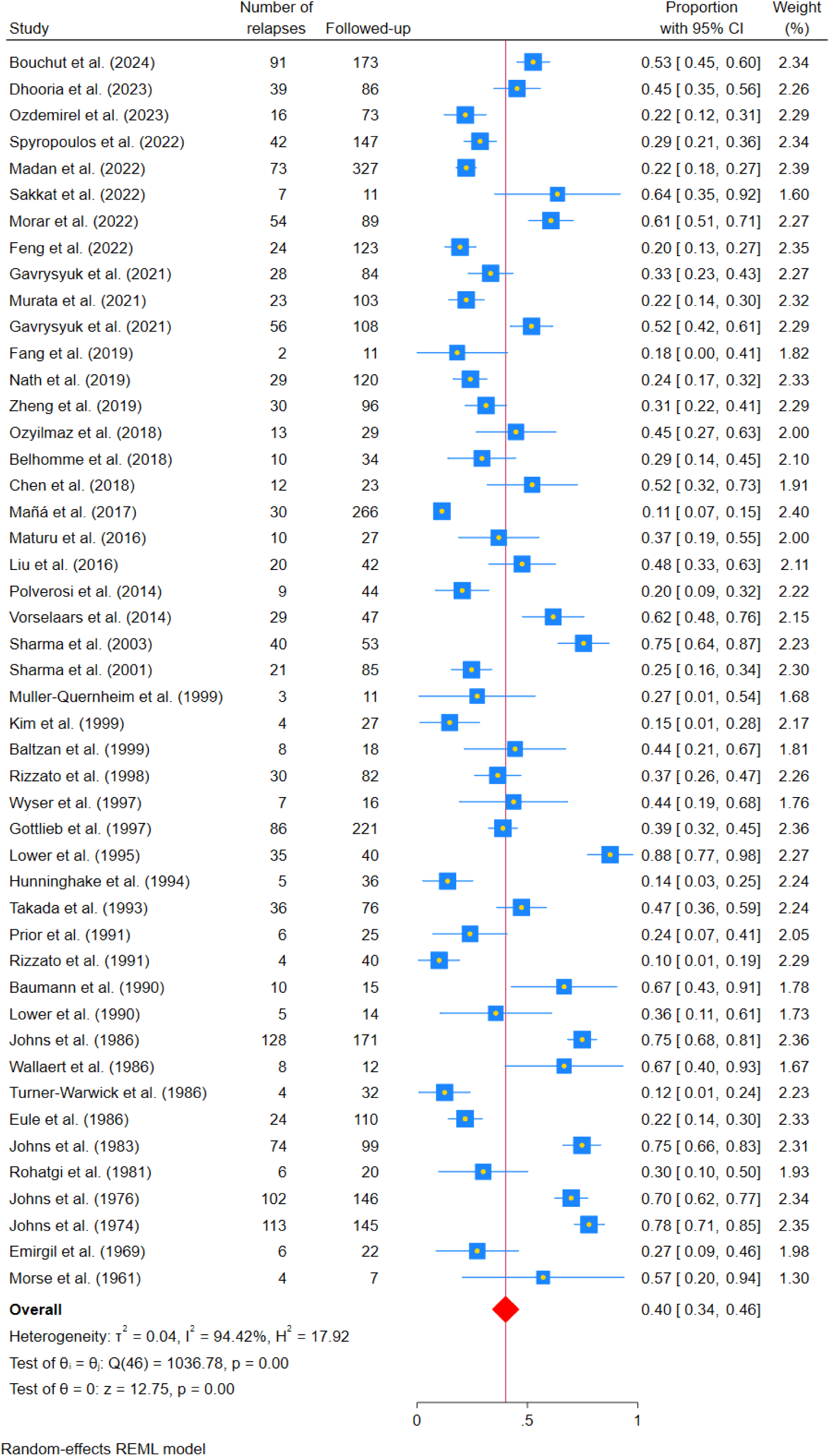
Pooled relapse prevalence in studies with only adult population.

**Figure S15:**
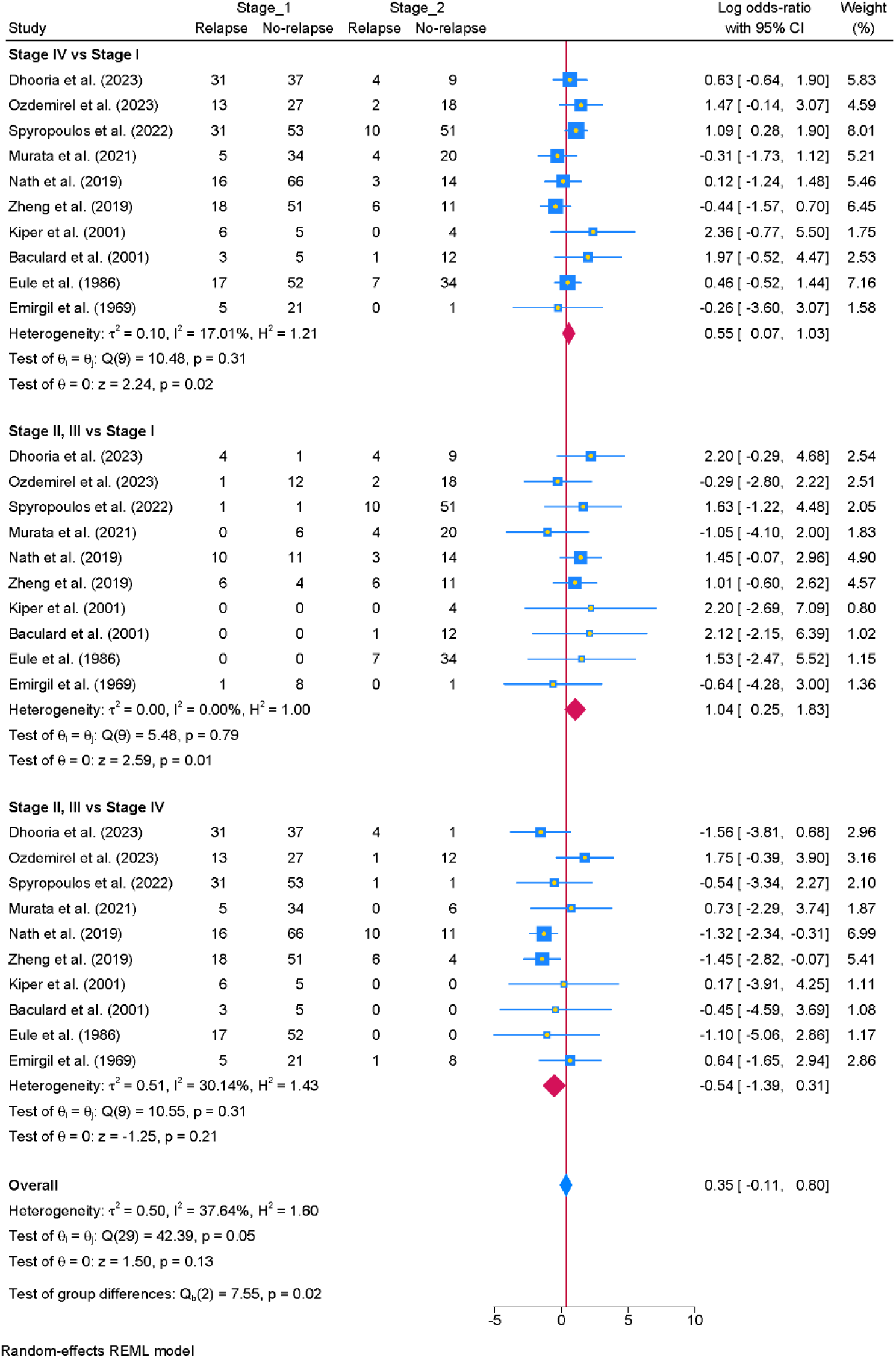
Forest plot comparing relapse rates across radiographic stages. Stage IV vs Stage I showed increased odds of relapse (log OR 0.55, p=0.02), while Stage II-III vs Stage I also showed higher odds (log OR 1.04, p=0.01). Stage II-III vs IV showed no significant difference (log OR −0.54, p=0.21).

**Figure S16:**
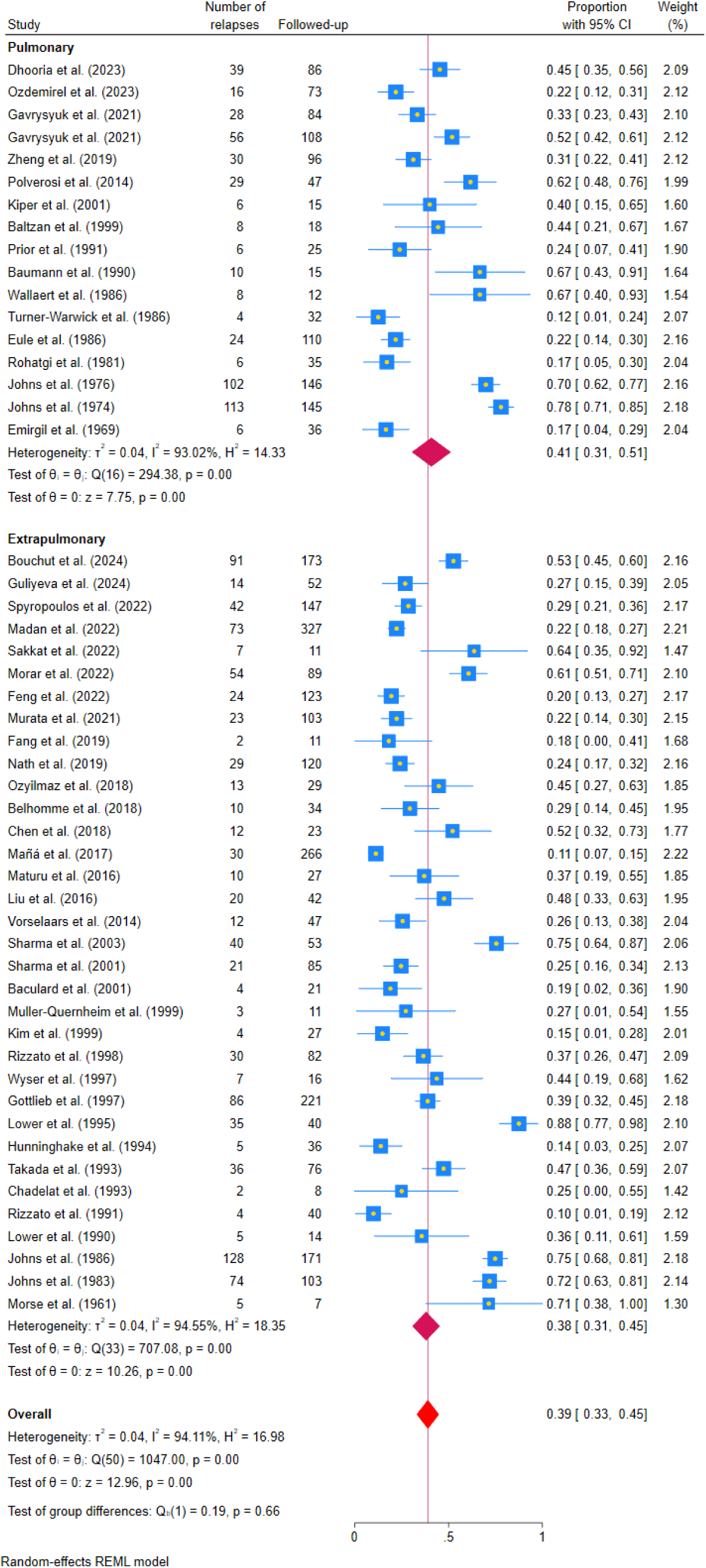
Forest plots of pooled relapse prevalence stratified by disease extent (isolated pulmonary vs. multiorgan).

**Figure S17:**
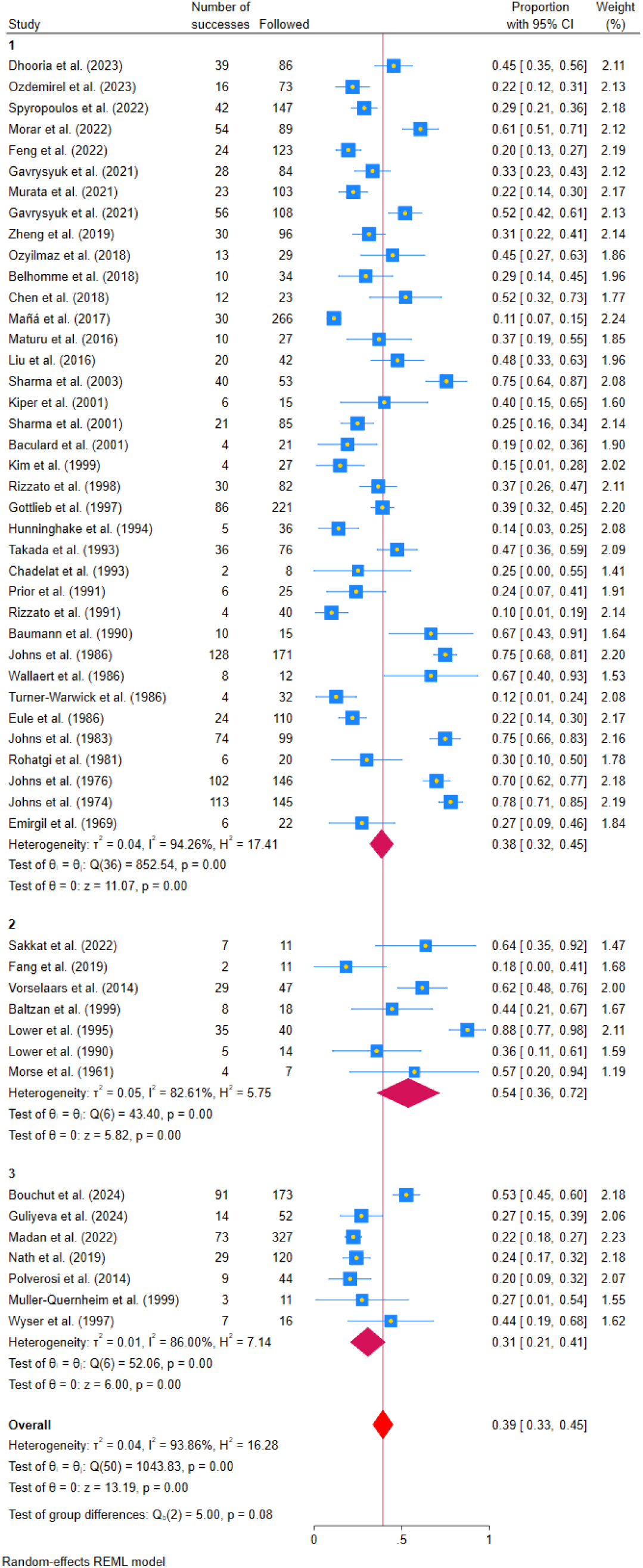
Forest plots of pooled relapse prevalence stratified by treatment type (corticosteroids, steroid-sparing agents, mixed).

**Figure S18:**
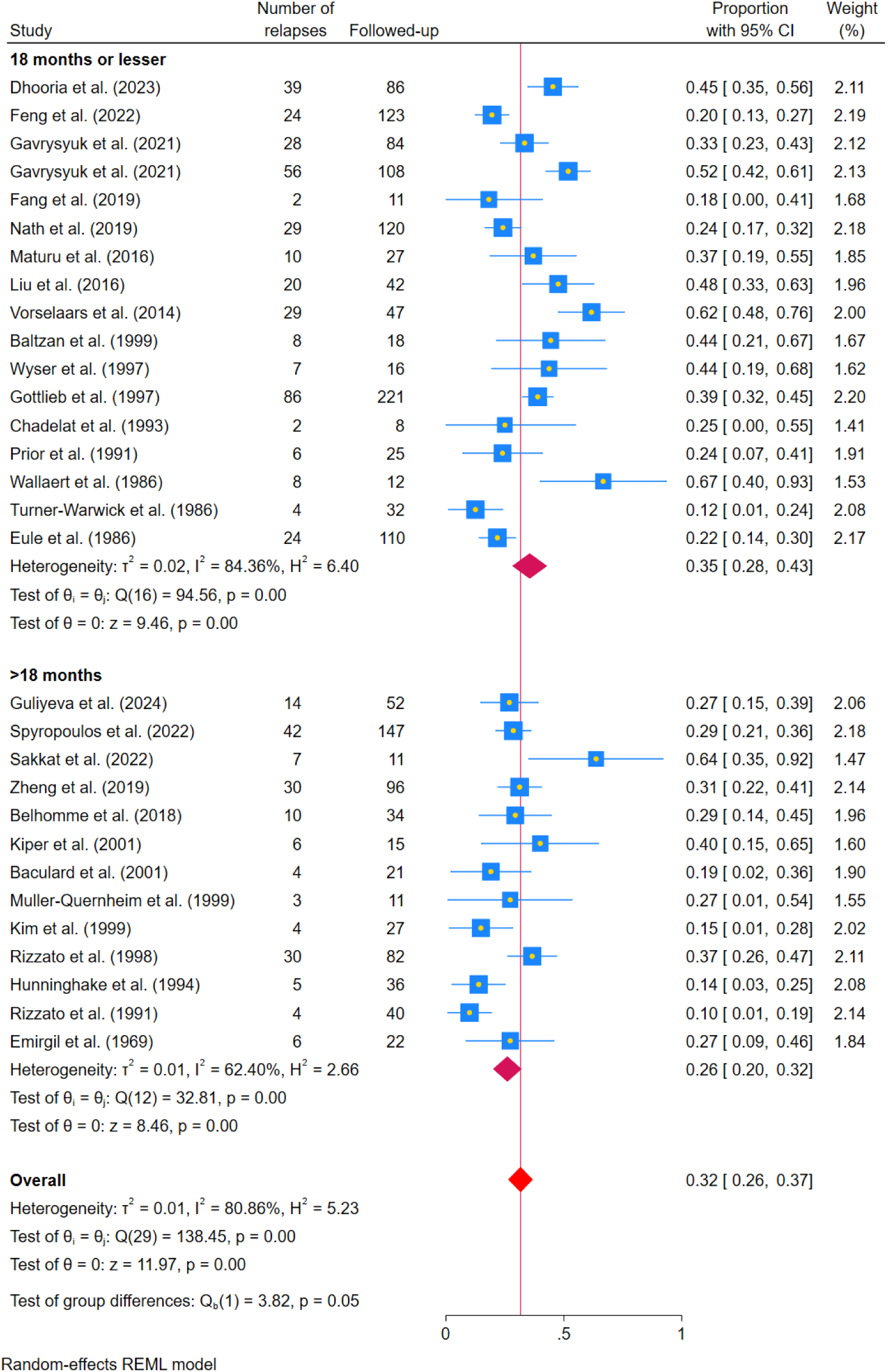
Subgroup analysis of relapse proportions in pulmonary sarcoidosis stratified by duration of corticosteroid therapy.

**Figure S19:**
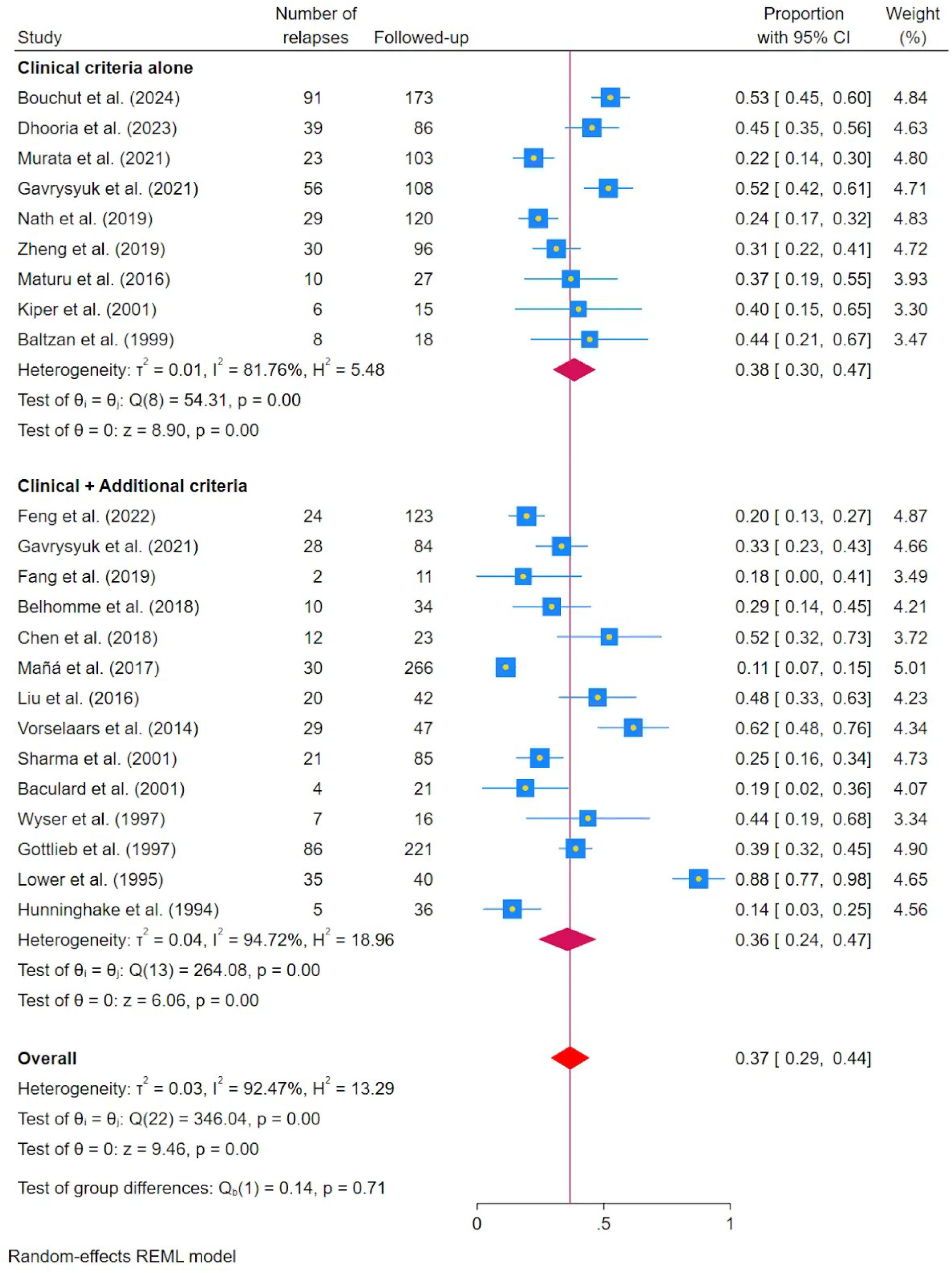
Effect of relapse definition criteria: clinical + additional criteria (radiological, functional, biomarkers) versus clinical criteria alone.

## SUPPLEMENTARY METHODS

### SM-1. Introduction

This document provides procedural justification for three post hoc methodological decisions: (i) extraction and operationalisation of corticosteroid therapy duration; (ii) rationale for classifying specific studies as NR (not reported) for duration; and (iii) the sensitivity analysis addressing the conceptual distinction between treatment failure and relapse on withdrawal. These sections respond directly to Major Comment #5 of Reviewer #3.

### SM-2. Duration of Therapy: Extraction Rules and Definitions

#### SM-2.1 Operational Definition

Duration of therapy was defined as the total calendar period of pharmacological therapy, expressed as the mean (or median, if mean unavailable) number of months from treatment initiation to cessation.

Duration was **never** equated with follow-up duration when both were reported.

**Six standardised extraction rules were applied:**

| Scenario | Rule | Example |
| --- | --- | --- |
| Single cohort duration reported | Record directly | Dhooria 2023: 6 months |
| Separate relapse/non-relapse data | Calculate weighted mean by subgroup n | Zheng 2019: 25.7 m (n=30) + 18.4 m (n=66) → 20.7 m |
| Range only, no mean/median | Arithmetic midpoint of range | Vorselaars 2014: 11–16 m → 13.5 m |
| Duration in weeks | Divide by 4.33 | Fang 2019: 53.1 wk ÷ 4.33 = 12.3 m |
| Duration in days | Divide by 30.44 | Rizzato 1991: 958 days ÷ 30.44 = 31.5 m |
| Follow-up only, strong proxy evidence | Record as proxy; note uncertainty | Kim 1999: 21 m follow-up used (CS withdrawn at end) |

#### SM-2.2 Data Extraction and Summary

Duration data were extracted independently by two reviewers from full text, methods, results, tables, and supplementary appendices. Discrepancies were resolved by consensus. Duration was coded NR when no estimable value could be derived after full-text review.

Of 51 included studies, duration was extractable from **30 studies (58.8%)**. Duration ranged from 1.5 months (Wallaert 1986, IV pulse) to 96 months (Johns 1986, chronic cohort); median 19.8 months (IQR 12.5–27.8 months).

### SM-3. Rationale for NR Classification by Study

#### SM-3.1 Johns Hopkins Series (Johns 1974, 1976, 1983, 1986) — Partial Inclusion

These four publications collectively contributed 590 treated patients but reported treatment duration as categorical distributions, not continuous means. Weighted means were estimated using interval midpoints — a validated imputation approach.

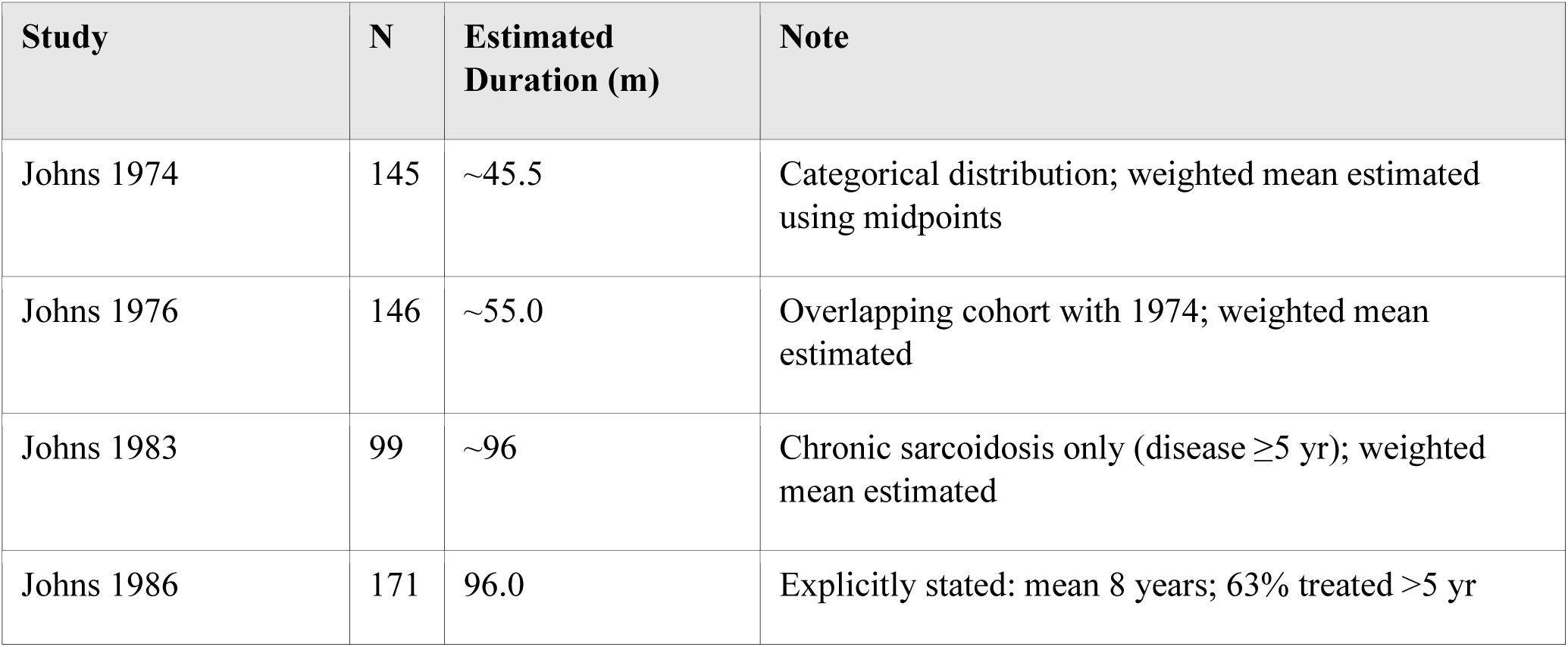

All four studies were retained in Model B to avoid disproportionate loss of long-duration data. Sensitivity analysis excluding the Johns series did not materially alter meta-regression coefficients (SM-5.3).

#### SM-3.2 Other Studies Coded NR — Summary

The 21 studies coded NR fell into five categories:

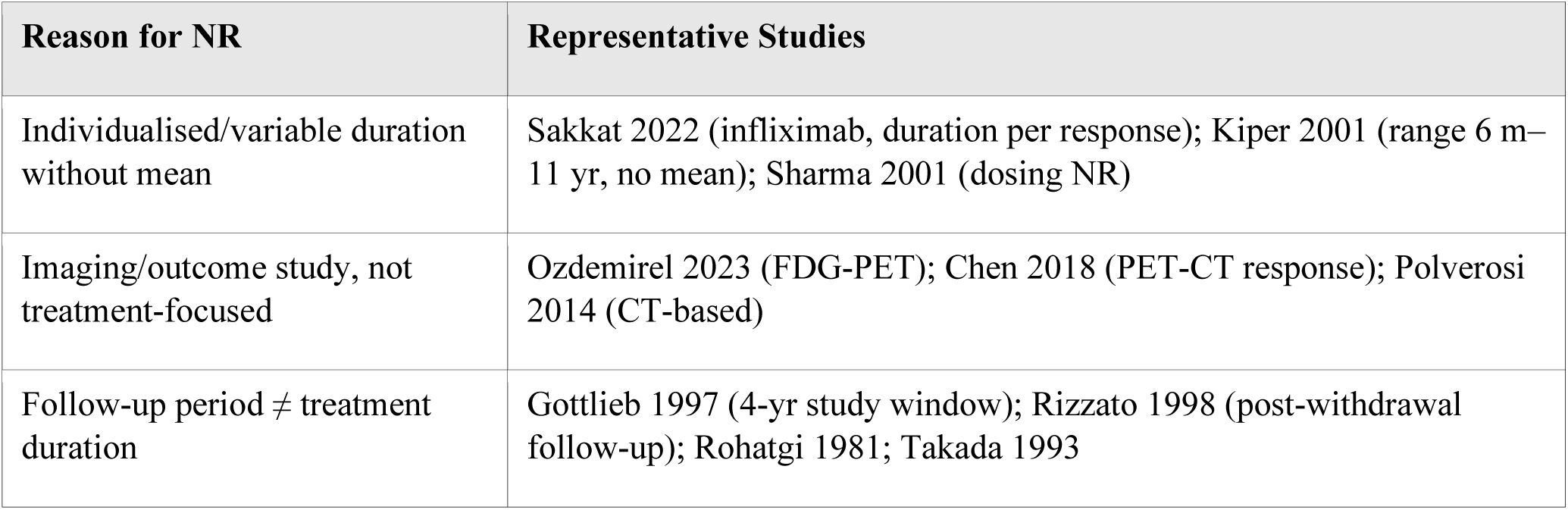
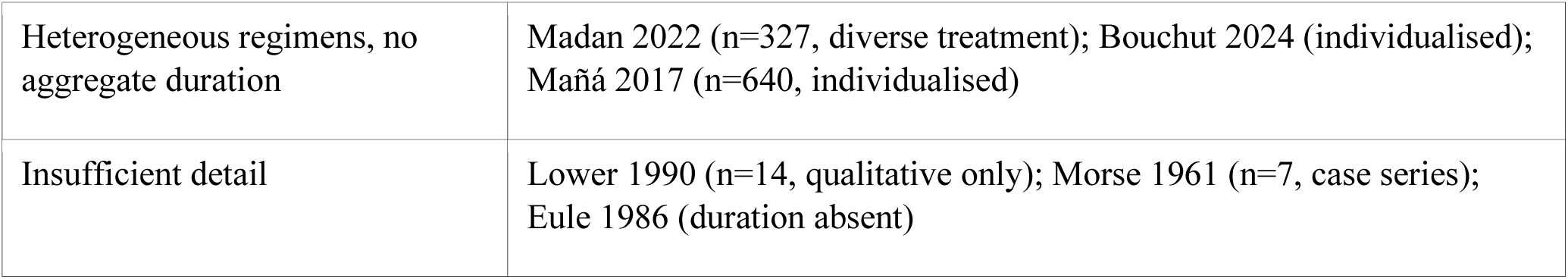

### SM-4. Treatment Failure vs Relapse on Withdrawal: Definition and Sensitivity Analysis

#### SM-4.1 Conceptual Distinction

Two distinct clinical phenomena are frequently conflated in the literature:

|  | Treatment Failure (Early Relapse) | Relapse on Withdrawal |
| --- | --- | --- |
| Definition | Disease re-emergence during or within 1 month of tapering/cessation, without a documented sustained remission interval | Recurrence of disease after documented remission, occurring on dose reduction or after cessation |
| Timing | During active treatment phase | After completion of, or during tapering of, treatment course |
| Clinical implication | Regimen inadequacy; escalation needed | Treatment dependency; reinstitution may be required |
| Post-2015 ERS/ATS term | Refractory disease / treatment failure | Relapse (current preferred term) |

#### SM-4.2 Primary Analysis Approach

The primary meta-analysis adopted an inclusive definition encompassing both categories, for three pre-specified reasons:

- The PROSPERO protocol (CRD420251075384) pre-specified inclusion of studies using any validated relapse definition, without restriction to post-withdrawal events, to maximise evidence breadth.
- A substantial proportion of pre-2005 studies did not explicitly distinguish between on-treatment failure and post-withdrawal relapse; excluding these would have resulted in >30% dataset attrition.
- The clinical question — overall burden of relapse in pharmacologically treated pulmonary sarcoidosis — is best served by an inclusive definition capturing both outcome types.

#### SM-4.3 Study Classification for Sensitivity Analysis

Each of the 51 studies was classified by two independent reviewers into one of three categories:

| Category | k (studies) | Criteria | Representative Studies |
| --- | --- | --- | --- |
| A — Pure relapse | 19 | Relapse explicitly post-withdrawal or post-taper; time-to-relapse referenced to cessation | Kim 1999, Muller-Quernheim 1999, Prior 1991, Wyser 1997, Baltzan 1999, Rizzato 1991/1998, Chadelat 1993, Hunninghake 1994, Dhooria 2023, Gavrysyuk 2021a/b, Zheng 2019, Maturu 2016, Nath 2019, Vorselaars 2014, Kiper 2001, Baculard 2001 |
| B — Mixed/unclear | 30 | Definition not restricted to post-withdrawal events, or not clearly specified; older studies using generic 'relapse' | Bouchut 2024, Guliyeva 2024, Spyropoulos 2022, Madan 2022, Morar 2022, Feng 2022, Fang 2019, Mañá 2017, Liu 2016, Johns 1974–1986, and others |
| C — Treatment failure | 2 | Primary outcome = on-treatment worsening or failure to achieve remission, without documented prior remission | Sakkat 2022 (infliximab arm); Liu 2016 (retreatment arm) |

Restriction to Category A studies (k=19) confirmed that the primary 39% pooled estimate was not materially inflated by inclusion of mixed-definition studies.

### SM-5. Sensitivity Analyses for Duration Meta-regression (Model B)

The primary Model B (7 covariates; n=30 studies) yielded a non-significant positive association between treatment duration and logit-relapse proportion (β = +0.021, 95% CI −0.004 to +0.047; p=0.103; adjusted R²=33.8% vs 18.7% for Model A). Three pre-specified sensitivity analyses were conducted:

| Analysis | Specification | Purpose |
| --- | --- | --- |
| Excl. estimated durations | k=23 (directly reported values only; excludes Johns 1974/76/83, Sharma 2003, Feng 2022, Belhomme 2018, Baculard 2001) | Confirms result is not an artefact of imputation |
| Excl. Johns Hopkins series | n=26 (removes four outlier studies with estimated durations 45–96 m vs median 19.8 m) | Tests for undue leverage of long-duration outliers |
| Categorical threshold analysis | Strata: $<12$ m / $12-<18$ m / $18-<24$ m / $\geq 24$ m (ERS/BTS/ATS guideline-aligned) | Examines non-linear effects; clinically interpretable between-group comparisons |

### SM-6. Key Limitations of Duration and Relapse-Type Analyses

| Limitation | Impact | Mitigation |
| --- | --- | --- |
| 21/51 studies (41.2%) did not report treatment duration | Model B restricted to 30 studies; potential selection bias | Baseline characteristics of duration-reported vs NR groups did not differ significantly in design, region, or relapse rate. Model A (n=51) serves as primary meta-regression. |
| Follow-up and treatment duration conflated in older studies | Risk of overestimating true treatment duration | Conservative extraction: duration coded NR when not clearly distinguished from surveillance. Follow-up retained as separate covariate. |
| Heterogeneity in treatment agent (CS vs SSA vs biologic) | Duration coefficient uninterpretable in isolation; longer studies may use different agents | Treatment-type subgroup analysis conducted separately. |
| Disease severity is an unmeasured confounder of both duration and relapse | Positive confounding may inflate duration coefficient (sicker patients treated longer, more likely to relapse) | Acknowledged in Discussion. Cannot be adjusted without individual patient data (IPD). |
| Definitional heterogeneity (treatment failure vs post-withdrawal relapse) | Up to ~30% of studies may include on-treatment failure in relapse counts | Pre-specified sensitivity analysis (SM-4) restricting to Category A studies did not materially change the pooled estimate. |
| 7 studies used estimated duration values | Introduces measurement uncertainty in the duration covariate | Sensitivity analysis excluding estimated values (SM-5) confirms results are not driven by these estimates. |

## REFERENCES

1. Baughman RP, Culver DA, Judson MA. A concise review of pulmonary sarcoidosis. Am J Respir Crit Care Med. 2011;183(5):573–581.

2. Valeyre D, Prasse A, Nunes H, Uzunhan Y, Brillet PY, Müller-Quernheim J. Sarcoidosis. Lancet. 2014;383(9923):1155–1167.

3. Dhooria S, Agarwal R, Sehgal IS, et al. High-dose prednisolone vs low-dose prednisolone in the management of pulmonary sarcoidosis (PREDMETH). Chest. 2023;163(6):1377–1388.

4. Vorselaars AD, Wuyts WA, Vorselaars VM, et al. Methotrexate vs azathioprine in second-line therapy of sarcoidosis. Chest. 2013;144(3):805–812.

5. Baughman RP, Costabel U, du Bois RM. Treatment of sarcoidosis. Clin Chest Med. 2008;29(3):533–548.

6. Paramothayan NS, Lasserson TJ, Jones PW. Corticosteroids for pulmonary sarcoidosis. Cochrane Database Syst Rev. 2005;(2):CD001114.

7. Page MJ, McKenzie JE, Bossuyt PM, et al. The PRISMA 2020 statement: an updated guideline for reporting systematic reviews. BMJ. 2021;372:n71.

8. Stroup DF, Berlin JA, Morton SC, et al. Meta-analysis of observational studies in epidemiology (MOOSE): a proposal for reporting. JAMA. 2000;283(15):2008–2012.

9. Hoy D, Brooks P, Woolf A, et al. Assessing risk of bias in prevalence studies: modification of an existing tool and evidence of interrater agreement. BMC Med Res Methodol. 2012;12:72.

10. DerSimonian R, Laird N. Meta-analysis in clinical trials. Control Clin Trials. 1986;7(3):177–188.

11. Egger M, Davey Smith G, Schneider M, Minder C. Bias in meta-analysis detected by a simple, graphical test. BMJ. 1997;315(7109):629–634.

12. Begg CB, Mazumdar M. Operating characteristics of a rank correlation test for publication bias. Biometrics. 1994;50(4):1088–1101.

13. Higgins JP, Thompson SG. Quantifying heterogeneity in a meta-analysis. Stat Med. 2002;21(11):1539–1558.

14. Baujat B, Mahé C, Pignon JP, Hill C. A graphical method for exploring heterogeneity in meta-analyses: application to a meta-analysis of 65 trials. Stat Med. 2002;21(18):2641–2652.

15. Galbraith RF. A note on graphical presentation of estimated odds ratios from several clinical trials. Stat Med. 1988;7(8):889–894.

16. Guyatt GH, Oxman AD, Vist GE, et al. GRADE: an emerging consensus on rating quality of evidence and strength of recommendations. BMJ. 2008;336(7650):924–926.

17. Judson MA, Costabel U, Drent M, et al. The WASOG sarcoidosis organ assessment instrument: an update of a previous clinical tool. Sarcoidosis Vasc Diffuse Lung Dis. 2014;31(1):19–27.

18. Baughman RP, Judson MA. Relapses of sarcoidosis: what are they and can we predict or prevent them? Eur Respir J. 2014;43(2):337–339.

19. Gottlieb JE, Israel HL, Steiner RM, Triolo J, Patrick H. Outcome in sarcoidosis: the relationship of relapse to corticosteroid therapy. Chest. 1997;111(3):623–631.

20. Soto-Gomez N, Peters JI, Nambiar AM. Diagnosis and management of sarcoidosis. Am Fam Physician. 2016;93(10):840–848.

21. Belhomme N, Jouneau S, Brinchault G, et al. Predictive factors for sarcoidosis relapse. Medicine (Baltimore). 2018;97(51):e13835.

22. Gavrysyuk VD, Merenkova IN, Dziublyk YA, et al. Efficacy and tolerability of methotrexate and methylprednisolone in pulmonary sarcoidosis. Diagnostics. 2021.

23. Ozdemirel TS, Ozkan C, Aydin F, et al. Predictors of relapse in pulmonary sarcoidosis. Turk Thorac J. 2023;24:100–106.

24. Murata M, Ohira S, Nishida C, et al. Relapse of cardiac sarcoidosis after treatment cessation. J Cardiol. 2021.

25. Bouchut M, Launay D, Hachulla E, et al. Musculoskeletal manifestations and relapse in sarcoidosis. Rev Med Interne. 2024.

26. Spyropoulos D, Sioula O, Karamanis T, et al. Ocular and skin sarcoidosis: long-term outcomes. Respir Med. 2022.

27. Sakkat A, Ghofranian A, Barnawi H, et al. Neurosarcoidosis refractory to corticosteroids treated with infliximab. Can Respir J. 2022.

28. Moravan M, Segal BM. Treatment of CNS sarcoidosis with infliximab and mycophenolate mofetil. Neurology. 2009;72(4):337–340.

29. Wijsenbeek MS, Culver DA. Treatment of sarcoidosis. Clin Chest Med. 2015;36(4):751–767.

30. da Costa CH, Silva VL, Fabricio-Silva GM, et al. HLA in a cohort of Brazilian patients with sarcoidosis. Hum Immunol. 2013;74(10):1326–1332.

31. Mirsaeidi M, Machado RF, Schraufnagel D, Sweiss NJ, Baughman RP. Racial difference in sarcoidosis mortality in the United States. Chest. 2015;147(2):438–449.

32. Burke RR, Stone CH, Havstad S, Rybicki BA. Racial differences in sarcoidosis granuloma density. Lung. 2009;187(1):1–7.

33. Vis R, Mathijssen H, Keijsers RG, et al. Prednisone vs methotrexate in treatment-naive cardiac sarcoidosis. J Nucl Cardiol. 2023;30(4):1543–1553.

34. Maturu VN, Rayamajhi SJ, Agarwal R, et al. Role of serial F-18 FDG PET/CT scans in assessing treatment response and predicting relapses in patients with symptomatic sarcoidosis. Sarcoidosis Vasc Diffuse Lung Dis. 2016;33(4):372–380.

35. Harman NL, Gorst SL, Williamson PR, et al. SCOUT — sarcoidosis outcomes taskforce: a systematic review of outcomes in pulmonary sarcoidosis. Sarcoidosis Vasc Diffuse Lung Dis. 2021;38(3):e2021034.

36. Baughman RP, Judson MA, Beaumont JL, et al. Evaluating the minimal clinically important difference of the King’s Sarcoidosis Questionnaire. Ann Am Thorac Soc. 2021;18(3):477–485.

37. Guliyeva V, Demirkan FG, Yigit RE, et al. A clinical overview of paediatric sarcoidosis: multicentre experience from Turkey. Mod Rheumatol. 2024;34(3):639–645.

38. Madan K, Sryma PB, Pattnaik B, et al. Clinical profile of 327 patients with sarcoidosis in India: an ambispective cohort study in a tuberculosis endemic population. Lung India. 2022;39(1):51–57.

39. Morar R, Feldman C. Sarcoidosis in Johannesburg, South Africa: a retrospective study. Afr J Thorac Crit Care Med. 2022;28(4):150–156.

40. Feng H, Yan L, Zhao Y, Li Z, Kang J. Neutrophils in bronchoalveolar lavage fluid indicating the severity and relapse of pulmonary sarcoidosis. Front Med (Lausanne). 2021;8:787681.

41. Gavrysyuk V, Merenkova I, Dziublyk Y, et al. Efficacy and tolerability of methotrexate and methylprednisolone in a comparative assessment of the primary and long-term outcomes in patients with pulmonary sarcoidosis. Diagnostics. 2021.

42. Fang C, Zhang Q, Wang N, Jing X, Xu Z. Effectiveness and tolerability of methotrexate in pulmonary sarcoidosis: a single center real-world study. Sarcoidosis Vasc Diffuse Lung Dis. 2019;36(3):217–227.

43. Nath A, Hashim Z, Khan A, et al. Experience of sarcoidosis and factors predicting relapse at a tertiary care institute in North India. Indian J Rheumatol. 2019;14(4).

44. Zheng Y, Wang H, Xu Q, et al. Risk factors of relapse in pulmonary sarcoidosis treated with corticosteroids. Clin Rheumatol. 2019;38(7):1993–1999.

45. Ozyilmaz E, Ozturk OG, Durmaz A, et al. Early prediction of sarcoidosis prognosis with HLA typing: a 5-year follow-up study. Sarcoidosis Vasc Diffuse Lung Dis. 2018;35(3):184–191.

46. Chen H, Jin R, Wang Y, et al. The utility of 18F-FDG PET/CT for monitoring response and predicting prognosis after glucocorticoids therapy for sarcoidosis. Biomed Res Int. 2018;2018:1823710.

47. Mañá J, Rubio-Rivas M, Villalba N, et al. Multidisciplinary approach and long-term follow-up in a series of 640 consecutive patients with sarcoidosis: cohort study of a 40-year clinical experience at a tertiary referral center in Barcelona, Spain. Medicine. 2017;96(29).

48. Liu Y, Qiu L, Wang Y, et al. The circulating Treg/Th17 cell ratio is correlated with relapse and treatment response in pulmonary sarcoidosis patients after corticosteroid withdrawal. PLoS One. 2016;11(2):e0148207.

49. Polverosi R, Russo R, Coran A, et al. Typical and atypical pattern of pulmonary sarcoidosis at high-resolution CT: relation to clinical evolution and therapeutic procedures. Radiol Med. 2014;119(6):384–392.

50. Vorselaars AD, Verwoerd A, van Moorsel CH, et al. Prediction of relapse after discontinuation of infliximab therapy in severe sarcoidosis. Eur Respir J. 2014;43(2):602–609.

51. Sharma SK, Balamurugan A, Pandey RM, Saha PK, Mehra NK. Human leukocyte antigen-DR alleles influence the clinical course of pulmonary sarcoidosis in Asian Indians. Am J Respir Cell Mol Biol. 2003;29(2):225–231.

52. Kiper N, Anadol D, Ozcelik U, Gocmen A. Inhaled corticosteroids for maintenance treatment in childhood pulmonary sarcoidosis. Acta Paediatr. 2001;90(8):953–956.

53. Sharma SK, Mohan A, Guleria JS. Clinical characteristics, pulmonary function abnormalities and outcome of prednisolone treatment in 106 patients with sarcoidosis. J Assoc Physicians India. 2001;49:697–704.

54. Baculard A, Blanc N, Boule M, et al. Pulmonary sarcoidosis in children: a follow-up study. Eur Respir J. 2001;17(4):628–635.

55. Mulle r-Quernheim J, Kienast K, Held M, Pfeifer S, Costabel U. Treatment of chronic sarcoidosis with an azathioprine/prednisolone regimen. Eur Respir J. 1999;14(5):1117–1122.

56. Kim DS, Paik SH, Lim CM, et al. Value of ICAM-1 expression and soluble ICAM-1 level as a marker of activity in sarcoidosis. Chest. 1999;115(4):1059–1065.

57. Baltzan M, Mehta S, Kirkham TH, Cosio MG. Randomized trial of prolonged chloroquine therapy in advanced pulmonary sarcoidosis. Am J Respir Crit Care Med. 1999;160(1):192–197.

58. Rizzato G, Montemurro L, Colombo P. The late follow-up of chronic sarcoid patients previously treated with corticosteroids. Sarcoidosis Vasc Diffuse Lung Dis. 1998;15(1):52–58.

59. Wyser CP, van Schalkwyk EM, Alheit B, Bardin PG, Joubert JR. Treatment of progressive pulmonary sarcoidosis with cyclosporin A: a randomized controlled trial. Am J Respir Crit Care Med. 1997;156(5):1371–1376.

60. Lower EE, Baughman RP. Prolonged use of methotrexate for sarcoidosis. Arch Intern Med. 1995;155(8):846–851.

61. Hunninghake GW, Gilbert S, Pueringer R, et al. Outcome of the treatment for sarcoidosis. Am J Respir Crit Care Med. 1994;149(4 Pt 1):893–898.

62. Takada K, Ina Y, Noda M, et al. The clinical course and prognosis of patients with severe, moderate or mild sarcoidosis. J Clin Epidemiol. 1993;46(4):359–366.

63. Chadelat K, Baculard A, Grimfeld A, et al. Pulmonary sarcoidosis in children: serial evaluation of bronchoalveolar lavage cells during corticosteroid treatment. Pediatr Pulmonol. 1993;16(1):41–47.

64. Prior C, Haslam PL. Increased levels of serum interferon-gamma in pulmonary sarcoidosis and relationship with response to corticosteroid therapy. Am Rev Respir Dis. 1991;143(1):53–60.

65. Rizzato G, Fraioli P, Montemurro L. Long-term therapy with deflazacort in chronic sarcoidosis. Chest. 1991;99(2):301–309.

66. Baumann MH, Strange C, Sahn SA. Do chest radiographic findings reflect the clinical course of patients with sarcoidosis during corticosteroid withdrawal? AJR Am J Roentgenol. 1990;154(3):481–485.

67. Lower EE, Baughman RP. The use of low dose methotrexate in refractory sarcoidosis. Am J Med Sci. 1990;299(3):153–157.

68. Johns CJ, Schonfeld SA, Scott PP, Zachary JB, MacGregor MI. Longitudinal study of chronic sarcoidosis with low-dose maintenance corticosteroid therapy: outcome and complications. Ann N Y Acad Sci. 1986;465(1):702–712.

69. Wallaert B, Ramon P, Fournier EC, et al. High-dose methylprednisolone pulse therapy in sarcoidosis. Eur J Respir Dis. 1986;68(4):256–262.

70. Turner-Warwick M, McAllister W, Lawrence R, Britten A, Haslam PL. Corticosteroid treatment in pulmonary sarcoidosis: do serial lavage lymphocyte counts, serum ACE measurements, and gallium-67 scans help management? Thorax. 1986;41(12):903–913.

71. Eule H, Weinecke A, Roth I, Wuthe H. The possible influence of corticosteroid therapy on the natural course of pulmonary sarcoidosis: late results of a continuing clinical study. Ann N Y Acad Sci. 1986;465(1):695–701.

72. Johns CJ, Zachary JB, MacGregor MI, Curtis JL, Scott PP, Terry PB. The longitudinal study of chronic sarcoidosis. Trans Am Clin Climatol Assoc. 1983;94:173–181.

73. Rohatgi PK, Ryan JW, Lindeman P. Value of serial measurement of serum angiotensin converting enzyme in the management of sarcoidosis. Am J Med. 1981;70(1):44–50.

74. Johns CJ, Macgregor MI, Zachary JB, Ball WC. Extended experience in the long-term corticosteroid treatment of pulmonary sarcoidosis. Ann N Y Acad Sci. 1976;278(1):722–731.

75. Johns CJ, Zachary JB, Ball WC Jr. A ten-year study of corticosteroid treatment of pulmonary sarcoidosis. Johns Hopkins Med J. 1974;134(5):271–283.

76. Emirgil C, Sobol BJ, Williams MH Jr. Long-term study of pulmonary sarcoidosis: the effect of steroid therapy as evaluated by pulmonary function studies. J Chronic Dis. 1969;22(2):69–86.

77. Morse SI, Cohn ZA, Hirsch JG, Schaeder RW. The treatment of sarcoidosis with chloroquine. Am J Med. 1961;30:779–784.

